# Comparative Immunogenicity, Safety and Efficacy Profiles of four COVID-19 Vaccine types in healthy adults: Systematic Review cum Meta-analysis of Clinical Trial data

**DOI:** 10.1101/2023.08.10.23293964

**Authors:** Si Qi Yoong, Priyanka Bhowmik, Debprasad Dutta

## Abstract

Four principal types of authorised COVID-19 vaccines include inactivated whole-virus vaccines, protein subunit vaccines, viral-vector vaccines and nucleic acid (mRNA and DNA) vaccines. Despite numerous Randomised Controlled Trials (RCTs), comprehensive systematic review and comparative meta-analysis have not been performed to validate the immunogenicity, safety and efficacy of COVID-19 vaccines in the healthy adult population. We aim to fulfil this unmet void. We searched for peer-reviewed articles about RCTs of the COVID-19 vaccines on healthy adults (18-64 years) available in eight major bibliographic databases (PubMed, EMBASE, Web of Science, Cochrane Library, Scopus, ScienceDirect, POPLINE, HINARI) till August 28, 2022. The Risk of Bias (RoB) was assessed using the Cochrane RoB-2. Random effects meta-analysis was conducted by pooling dichotomous outcomes using risk ratios (safety outcomes) and continuous outcomes using standardised mean differences (immunogenicity outcomes). Efficacy outcomes were summarised narratively. Moderate to high-quality evidence suggests that those receiving COVID-19 vaccines had significantly higher immune responses compared to placebo. Serious adverse events were rare, confirming that COVID-19 vaccines were safe and immunogenic for the healthy adult population. Remarkably, adverse events were the least common in inactivated vaccines, and nucleic acid vaccines were the most immunogenic. The efficacies of COVID-19 vaccines ranged from 21.9% to 95.9% in preventing COVID-19. We endorse all four types of COVID-19 vaccines for public health policy implementing taskforces. Yet, meta-analyses based on individual patient data are warranted for more extensive measurement of differential impacts of COVID-19 vaccines on different genders, ethnicities, comorbidities and types of vaccine jabbed.

## 1. Introduction

The Coronavirus Disease 2019 (COVID-19) is an infectious respiratory communicable disease caused by Severe Acute Respiratory Syndrome Corona Virus 2 (SARS-CoV-2), originating in Wuhan, China, in early December 2019 [1]. World Health Organization (WHO) announced the outbreak as a global pandemic on March 11, 2020 [2]. COVID-19 is a systemic disease with both short-, intermediate- and long-term physical and mental health impacts [3, 4]. Majority of patients experience mild to moderate symptoms and 5–10% suffer from severe or debilitating disease. Therefore, the development of effective and safe vaccines and novel therapeutics is deemed a global exigency [5].

SARS-CoV-2 belongs to the genus *Betacoronavirus* under the *Coronaviridae* family and has four primary structural proteins, viz. Spike (S), Membrane (M) and Envelope (E) proteins in the viral surface, and Nucleocapsid (N) protein in the ribonucleoprotein core [6]. S proteins bind with a host cell receptor, angiotensin-converting enzyme 2 (ACE2), which is extensively expressed in pulmonary alveolar cells, cardiac myocytes, vascular endothelium and various other cell types, leading to viral invasion [7]. Most COVID-19 vaccines innovated so far have targeted the S protein. S protein consists of a membrane-distal S1 moiety and a membrane-proximal S2 moiety and presents on the viral envelope as a homotrimer (S1-S2 and S2′). The S1 subunit facilitates ACE2 recognition via its receptor-binding domain (RBD), whereas the S2 subunit enables membrane fusion during viral entry [8].

Four major types of COVID-19 vaccines are in clinical trials and/or have received emergency use authorisation globally: inactivated whole-virus vaccines, protein subunit vaccines, viral vector vaccines and nucleic acid (mRNA and DNA) vaccines. Inactivated whole-virus vaccine candidates contain attenuated SARS-CoV-2 viruses that induce immune responses similar to their real counterparts without causing disease. Protein subunit vaccines contain antigenic parts of the SARS-CoV-2 virus rather than the whole virus to trigger an immune response. Viral vector vaccines utilise modified viruses such as adenoviruses to deliver antigen-encoding genes which encode the surface spike proteins found on the virus and are delivered into human cells. Nucleic acid vaccines contain viral genetic material to provide immunity against the virus particles by encoding the viral antigen [9]. Vaccines offer protection against COVID-19 disease by eliciting both humoral and cellular immune responses [10], which work synergistically to ultimately induce neutralising antibodies crucial for virus clearance by targeting the S protein, thus preventing infection and risk reduction of severe COVID-19 disease [6, 11].

Meta-analyses on immunogenicity, safety and efficacy of COVID-19 vaccine trials among adults published till date have pooled trial data without differentiating between age groups and accounting for comorbidities [12–15], although these covariates markedly influence vaccine efficacy and immune response. With the rapid development of COVID-19 vaccine candidates, clinicians, policymakers and the public at large experienced confusion in deciding which vaccines/vaccine type would be more effective and which would be safer. A multitude of meta-analyses focused on patient groups with various comorbidities and in the younger population [16–20]. To the best of our knowledge, no meta-analysis has been conducted on the effects of COVID-19 vaccines in the healthy adult population. Due to the rapid development and publication of COVID-19 vaccine trial data, an updated systematic review and meta-analysis is needed. Hence, the current systematic review and meta-analysis aimed to compare the immunogenicity, safety, and efficacy of different types of COVID-19 vaccines in healthy adults.

## 2. Methods

We reported this systematic review and meta-analysis in accordance with the Preferred Reporting Items for Systematic Reviews and Meta-Analyses (PRISMA) [21]. The protocol was registered on PROSPERO (CRD42022314578).

### 2.1. Study selection criteria

We included peer-reviewed studies evaluating COVID-19 vaccine candidates irrespective of language and publication date. They must be randomised controlled trials (RCT) (Phase I-IV). Preclinical studies, and those of other study designs (e.g., quasi-experimental, reviews, opinion articles), publication types (e.g., conference abstracts, letters to editor etc.) and non-peer-reviewed articles (e.g., preprints, grey literature) were excluded.

Participants who were non-pregnant, non-lactating, healthy adults (18-64 years old) were included. When RCTs reported data on mixed populations, e.g., those with comorbidities or adults aged 65 years and above, we extracted data concerning only the subgroups of interest to our review. We excluded the trial if less than 90% of participants met the inclusion criteria (e.g., studies which mainly recruited participants aged <18 and >64 years old, >10% of participants had comorbidities which put them at risk of severe COVID-19 infection or immunosuppression, e.g., cancer, uncontrolled diabetes, cardiovascular disease, obesity). Given many vaccines are under development, this review focused on vaccines with potential clinical applicability; hence vaccines which ceased further development, or Phase I trials with very small sample sizes (with less than 20 participants in the intervention arm) were excluded unless the vaccine had been investigated in further trials.

In terms of intervention, all four types of COVID-19 vaccine candidates at any RCT phase (nucleic acid, viral vector, inactivated virus, and protein subunit vaccines) were eligible. Comparators were as defined by trials, which included placebo (e.g., saline, vaccine adjuvant or vaccine protecting for other diseases such as meningococcal conjugate vaccine) or no vaccine. However, studies on co-administering different vaccines were excluded, e.g., a COVID-19 vaccine and influenza vaccine.

Studies which evaluated at least one outcome (immunogenicity, safety and/or efficacy) were included in the review. Immunogenicity outcomes included humoral immunity [(geometric mean titres (GMT) and 95% confidence interval (CI)] of anti-RBD IgG, anti-S protein IgG and neutralising antibodies) and cell-mediated immunity (T-cell response). Safety outcomes of COVID-19 vaccine candidates included any adverse events, local, systemic, and serious adverse events. Efficacy outcomes included the number of COVID-19 infections, hospitalisations, ICU admissions, severe illness, and deaths due to COVID-19.

### 2.2. Search strategy

The detailed search strategy is presented in Supplementary File 1. We systematically searched 8 principal databases (PubMed, EMBASE, Web of Science, Cochrane Library, Scopus, ScienceDirect, POPLINE, HINARI) using keywords such as ‘safety’, ‘immunity’, ‘vaccine efficacy’ and “covid 19 vaccine’ for eligible articles on 18-19 April 2022. We also hand-searched the New England Journal of Medicine for relevant articles, as many COVID-19 vaccine RCTs were published in this journal. We searched trial registries (ClinicalTrials.gov, WHO International Clinical Trials Registry Platform) to ensure that all relevant published studies were included. Finally, reference lists of relevant studies and reviews were assessed. Initial search results were uploaded into EndNote X20, where duplicates were removed automatically and manually. Screening of titles and abstracts was done by PB and SQY using Rayyan (http://rayyan.qcri.org). They then independently assessed full texts for eligibility. Discrepancies were discussed until a consensus was reached. Given the rapid publication of COVID-19 vaccine trials, we checked regularly for peer-reviewed articles for relevant articles. The final cutoff date for inclusion into the review was August 28, 2022.

### 2.3. Data extraction

Data were extracted using a pre-piloted data extraction sheet by SQY and PB. Discrepancies were discussed until a consensus was reached. Information extracted includes author, year, country, study design, participant characteristics, vaccine characteristics, type of placebo, immunogenicity, safety, and efficacy outcomes.

### 2.4. Quality appraisal

Risk of bias (ROB) was independently assessed by PB and SQY for each study using the Revised Risk of Bias tool, and discrepancies were discussed until a consensus was reached [22]. ROB was assessed using 5 domains (bias arising from randomisation process, deviations from intended interventions, missing outcome data, outcome measurement, selection of reported results), and each domain was rated as ‘low risk of bias’, ‘high risk of bias’, or ‘some concerns’. We assessed deviations from interventions based on the effect of assignment to intervention (the intention-to-treat effect). Overall ROB for each study was evaluated accordingly, and ratings were visualised using Robvis [23].

The overall quality of evidence was rated following Grading of Recommendations, Assessment, Development and Evaluations (GRADE) guidelines and justifications were provided in Evidence Profile tables generated using GRADEproGDT software [24].

### 2.5. Synthesis approach

Meta-analyses were performed using Review Manager Version 5.4.1. The random-effects model was used for all analyses as it accounts for between-study heterogeneity. Meta-analysis was conducted only at timepoints which were investigated by 3 or more studies. For immunogenicity outcomes, standardised mean differences (SMD) of log-transformed geometric mean titers were selected as different assays were used, and that meta-analysis of skewed data can be performed using a natural log transformation [25, 26]. When geometric median titers were reported, we transformed them into geometric means using established formulas if possible [27]. For safety and efficacy outcomes, dichotomous data were pooled using risk ratios (RR) as the effect size. When meta-analysis was not possible (e.g., dissimilar outcomes, timepoints, inadequate data for meta-analysis, only descriptive/graphical data available), outcomes were summarised narratively.

Cochran’s Q test and I² statistics were used to evaluate heterogeneity. Statistically significant heterogeneity was set at p < 0.10. Heterogeneity was unimportant when I² = 0–40%, moderate when I² = 30–60%, substantial when I² = 50–90% and considerable when I² = 75–100%. If there were more than 10 studies in a meta-analysis and significant heterogeneity was found, subgroup and sensitivity analysis were used to investigate sources of heterogeneity [26]. We predefined subgroups to be based on age, sex and vaccine type (nucleic acid, viral vector, inactivated virus and protein subunit vaccines). There was a significant subgroup difference when p < 0.10 [28]. Sensitivity analysis was done by excluding each study. If results remain consistent, they were construed as robust. When results differed, they were treated with caution. If there were more than 10 studies in a meta-analysis, publication bias was assessed using visual inspection of funnel-plot asymmetry, Begg’s and Egger’s test [26] using Jamovi version 1.6.

## 3. Results

### 3.1. Search findings

The initial search yielded 20482 articles. After the removal of duplicates, 13112 articles were screened using titles and abstracts. Full texts of 113 articles were assessed, and finally, 41 RCTs were included in the systematic review [29–69]. The PRISMA diagram is shown in Figure 1.

**Fig 1.**
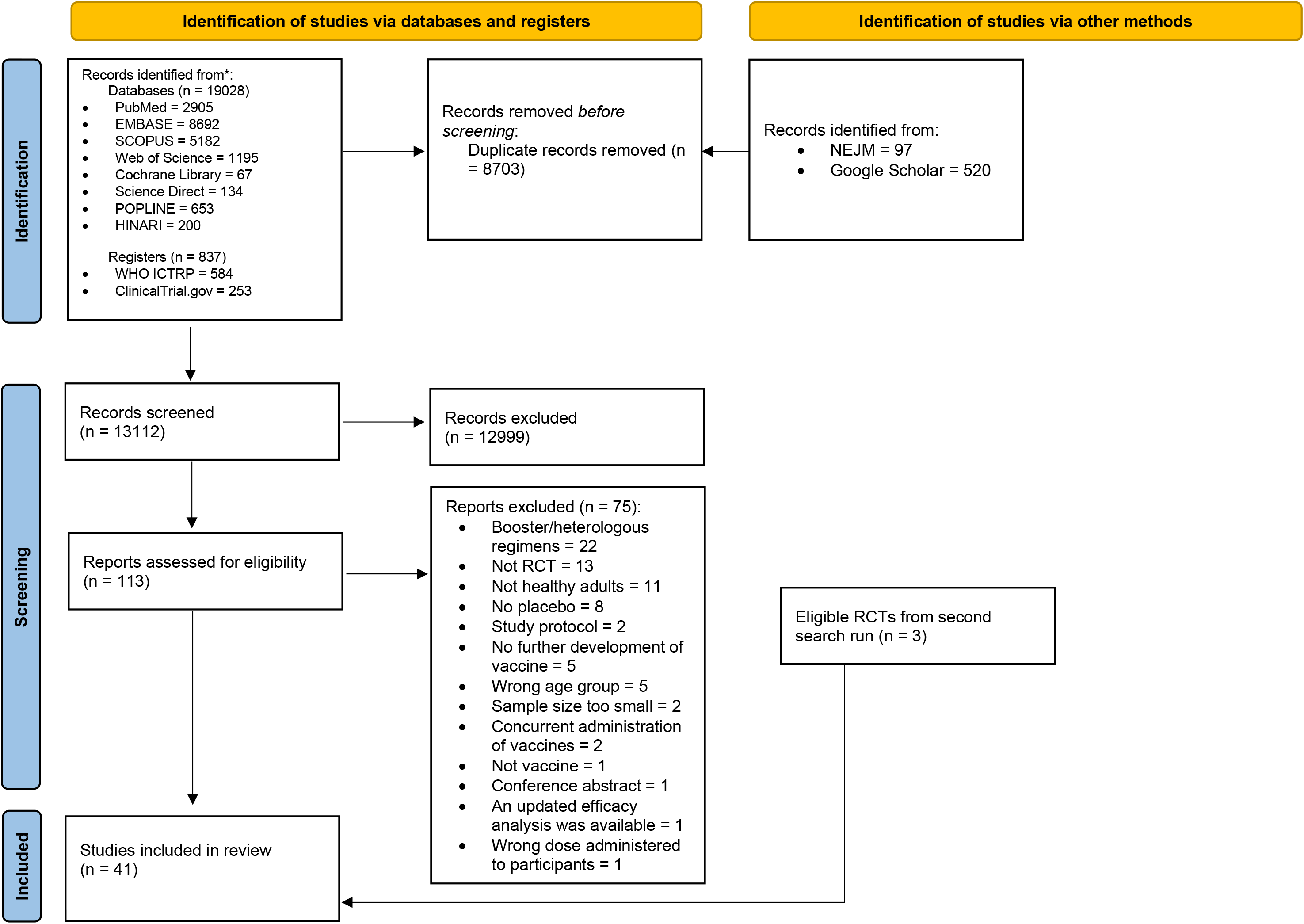
PRISMA diagram showing study selection process.

### 3.2. Characteristics of included studies

Studies were published from 2020 to 2022 across 25 countries, most commonly in China (n = 14), US (n = 8) and Japan (n = 5) (Table 1). Forty-one studies on 26 vaccines were included, of which 14 studies were on protein subunit vaccines, 12 on inactivated vaccines, 9 on viral vector vaccines and 6 on nucleic acid vaccines. Most were phase 1-2 RCTs, and there were 6 phase 3 RCTs [35, 42, 47, 54, 57, 65]. There was a total of 118 377 participants, with sample sizes ranging from 15 to 37594.

**Table 1.**
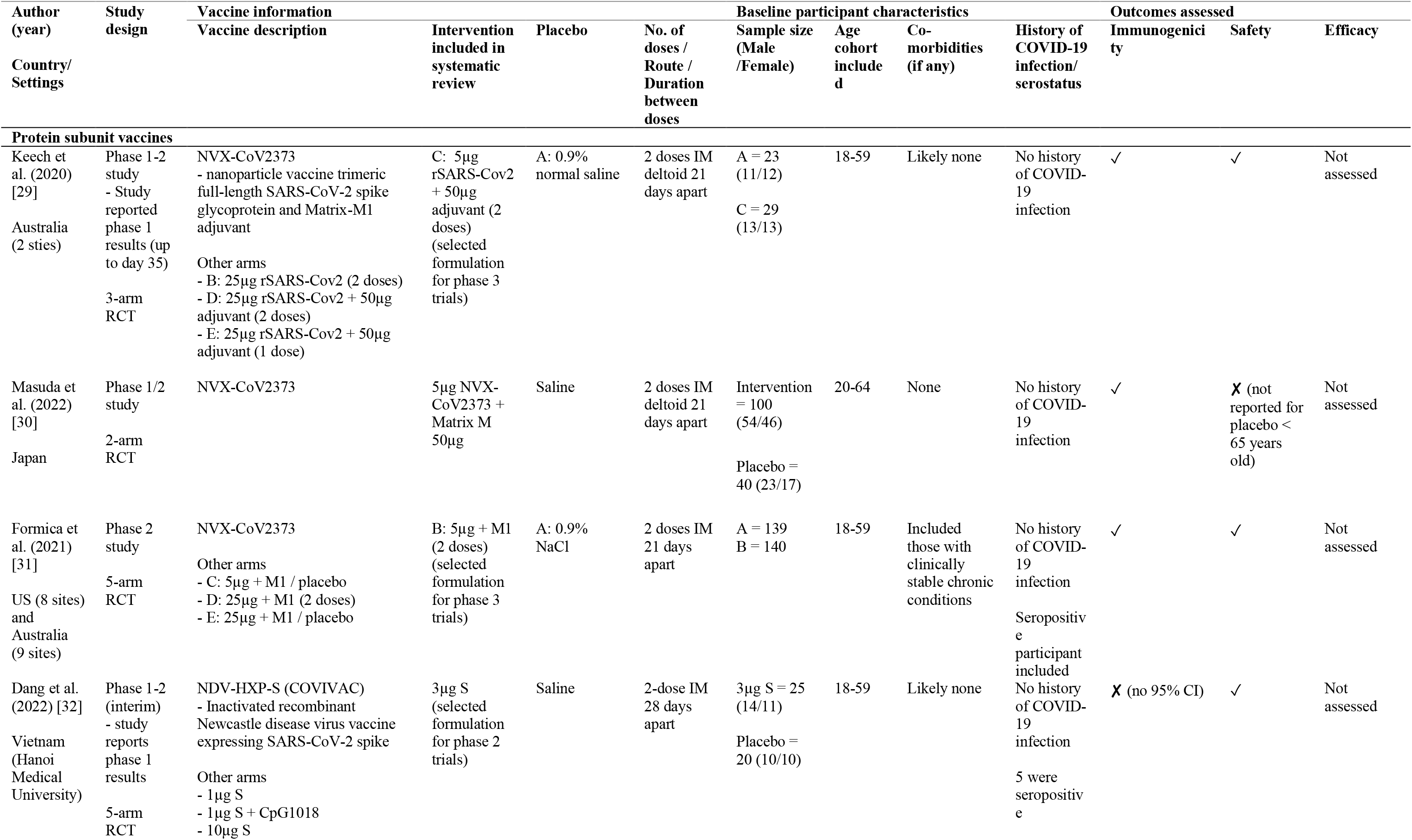
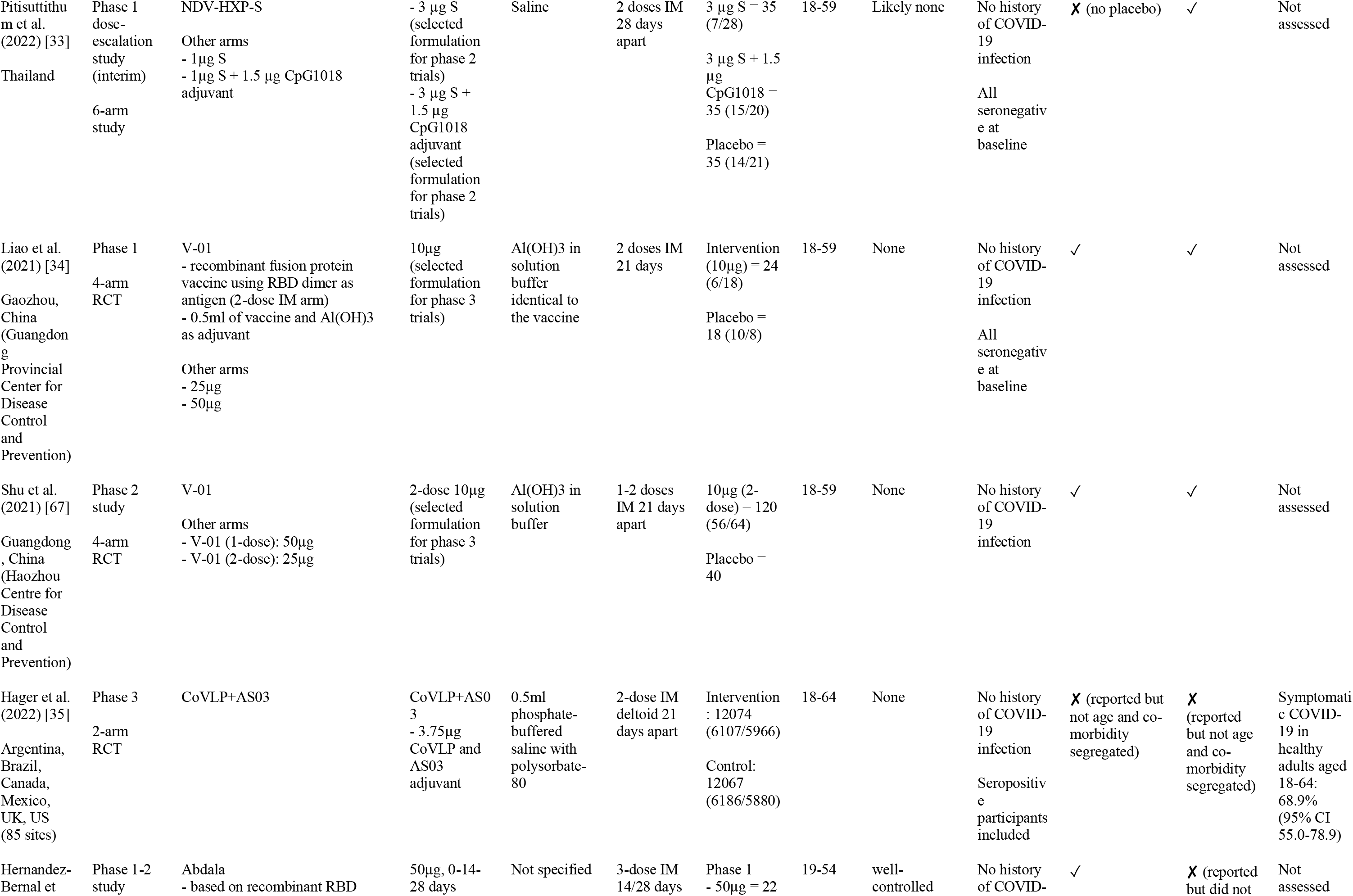
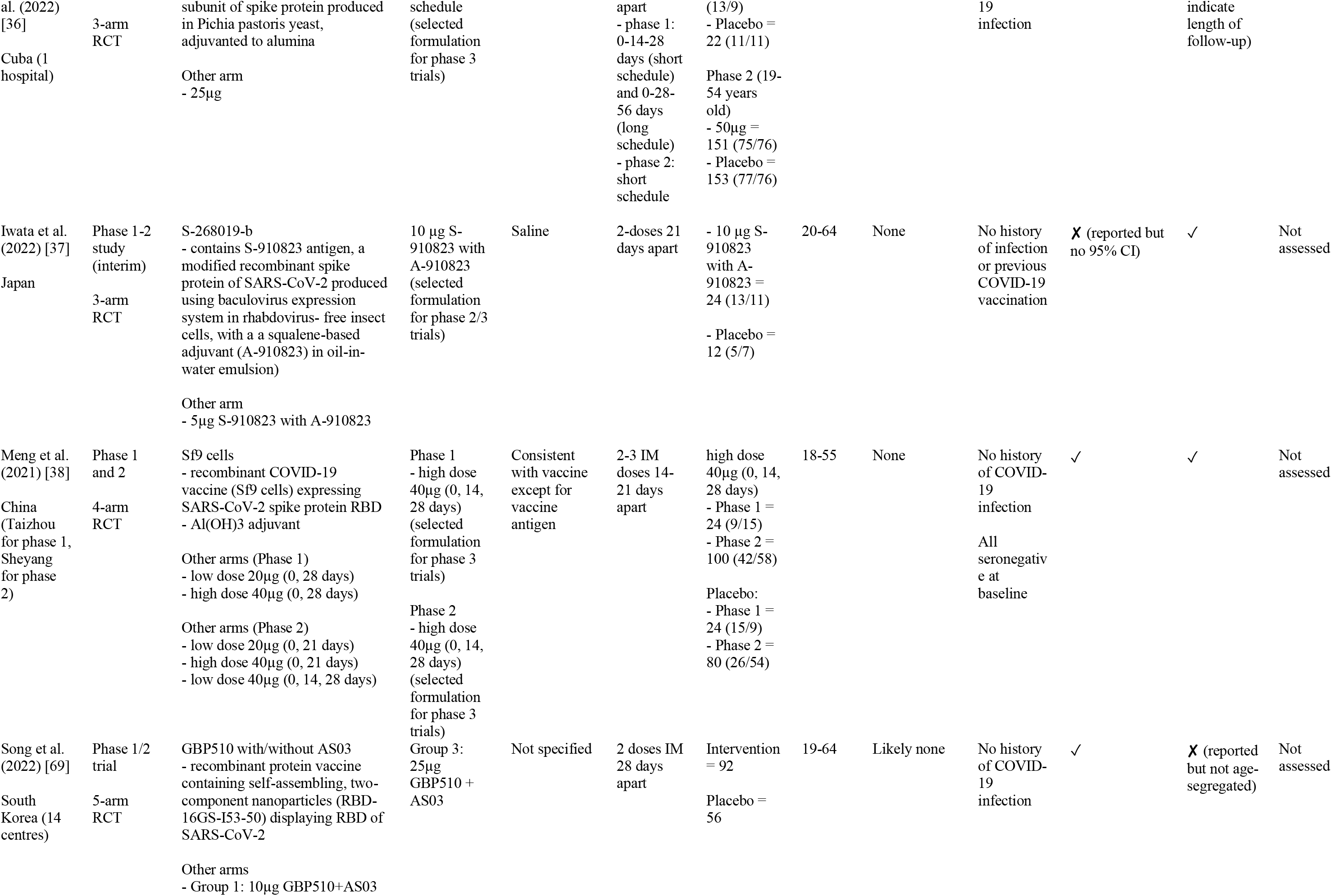
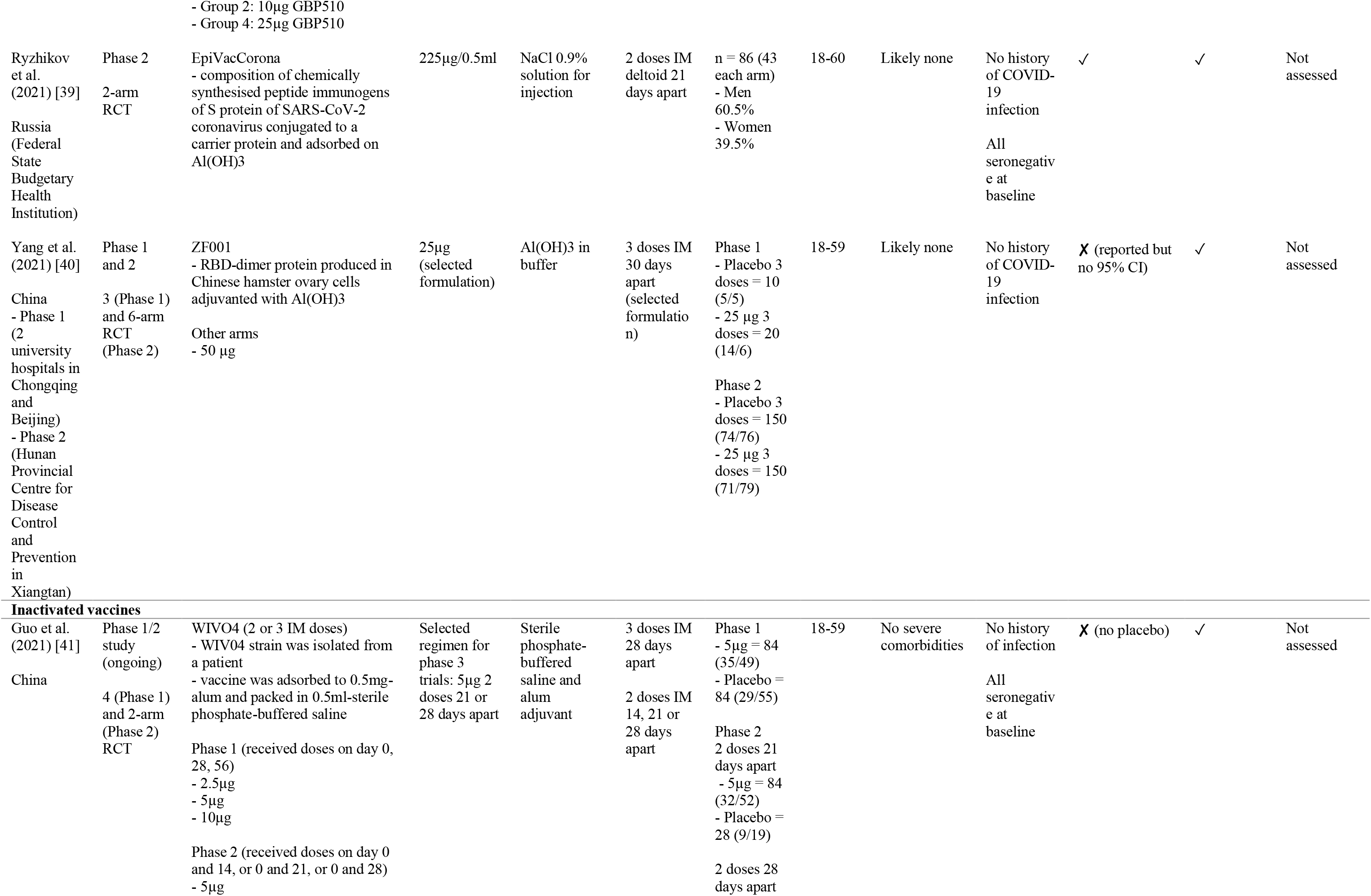
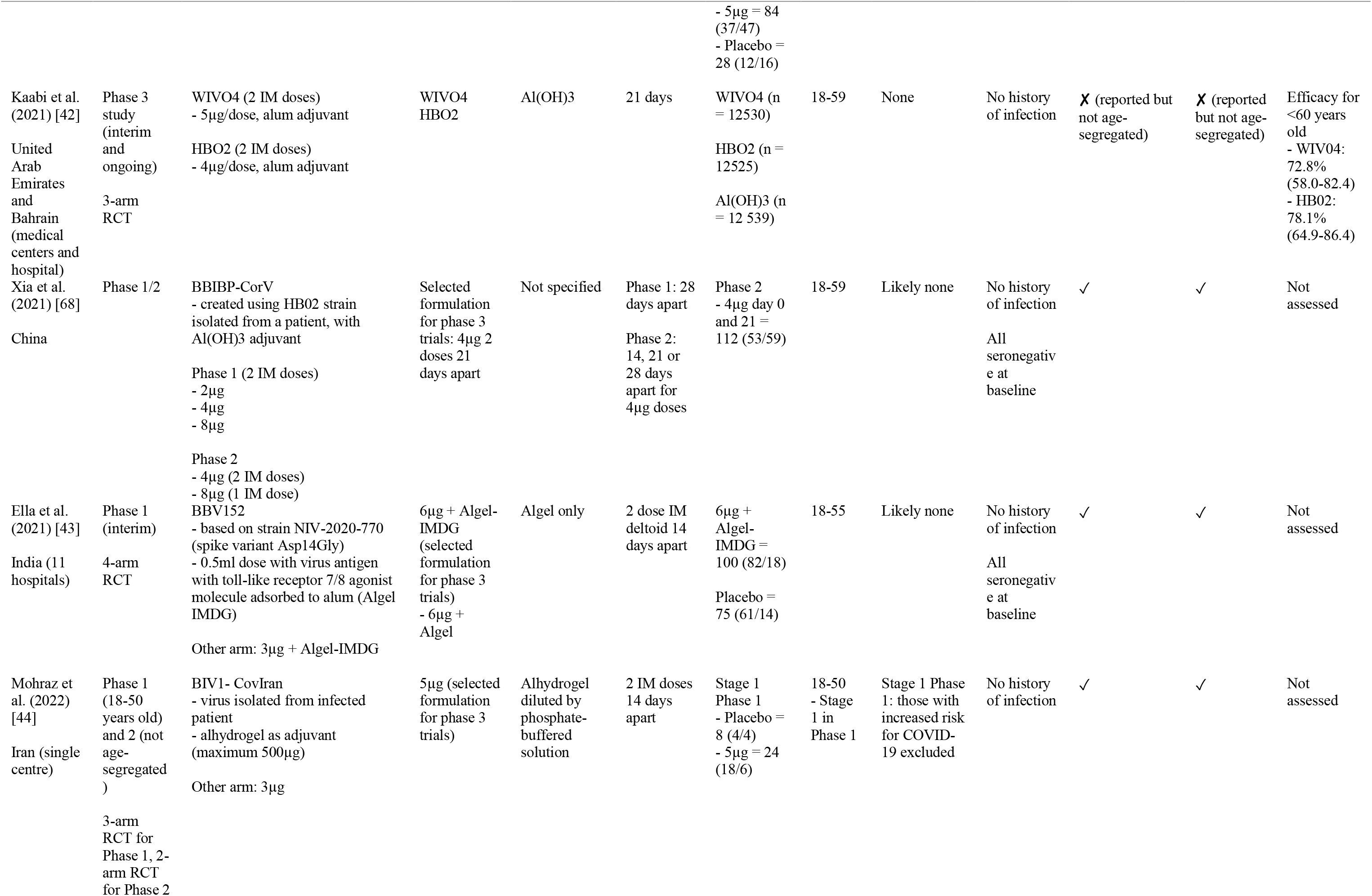
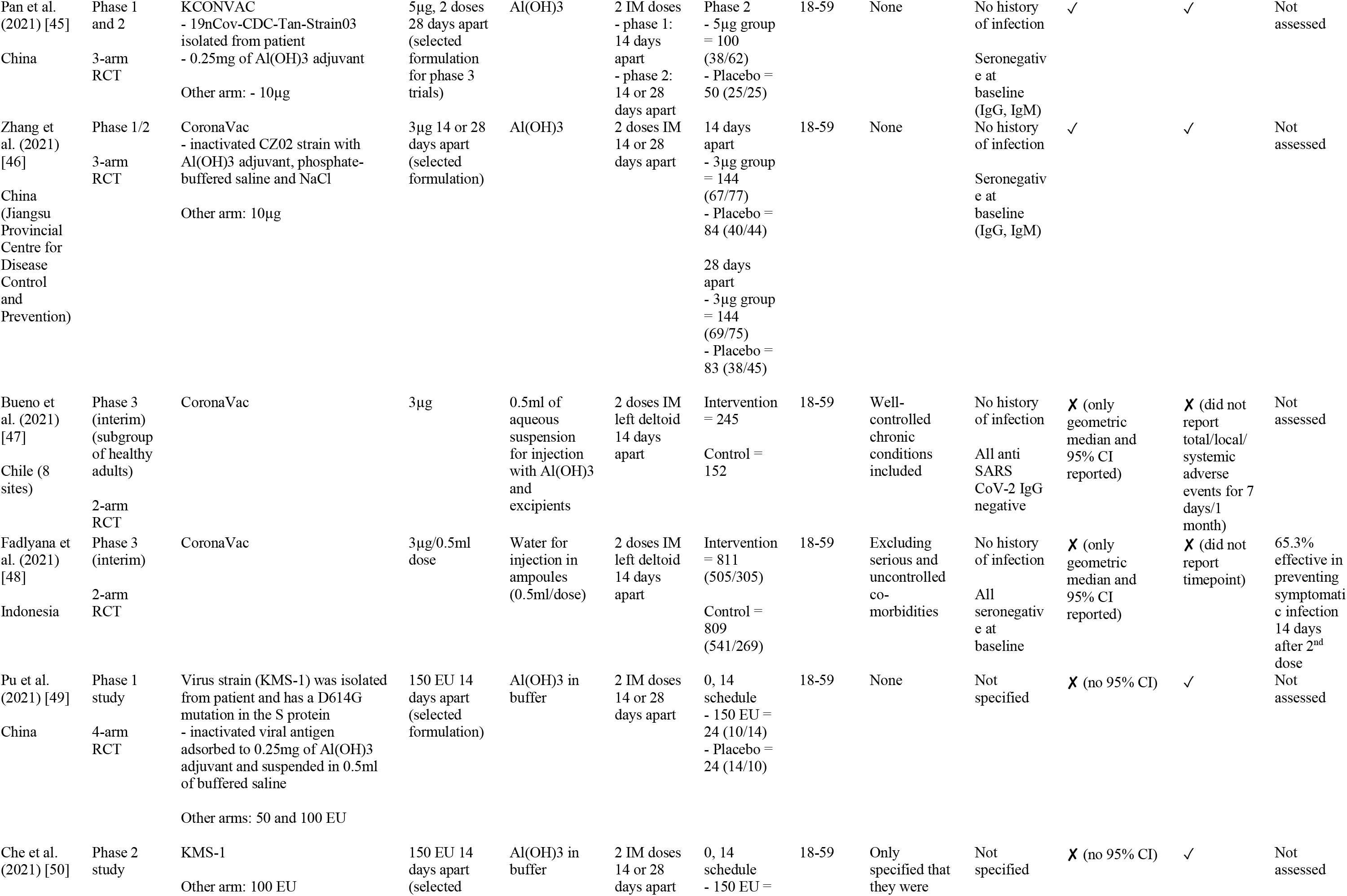
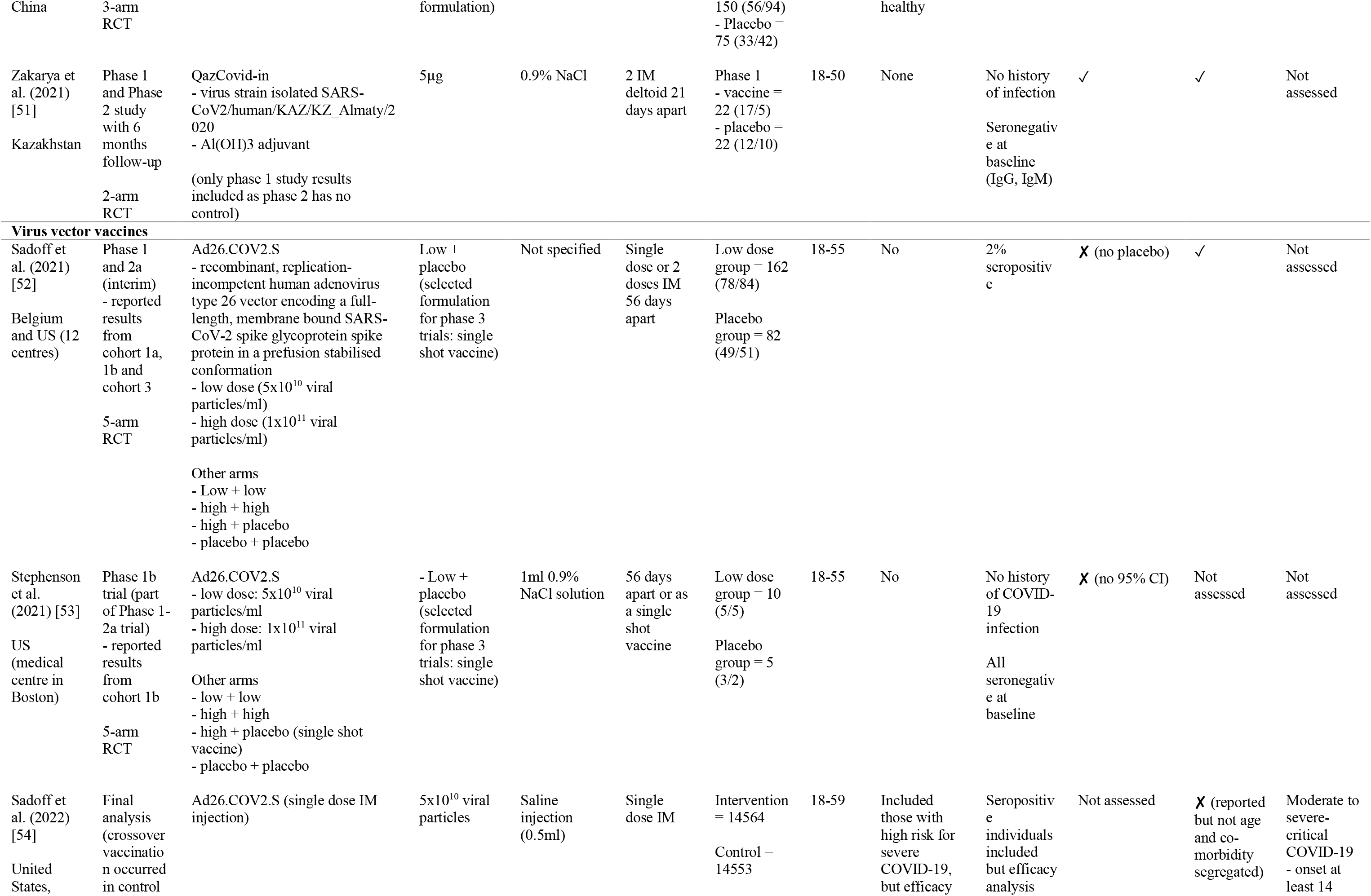
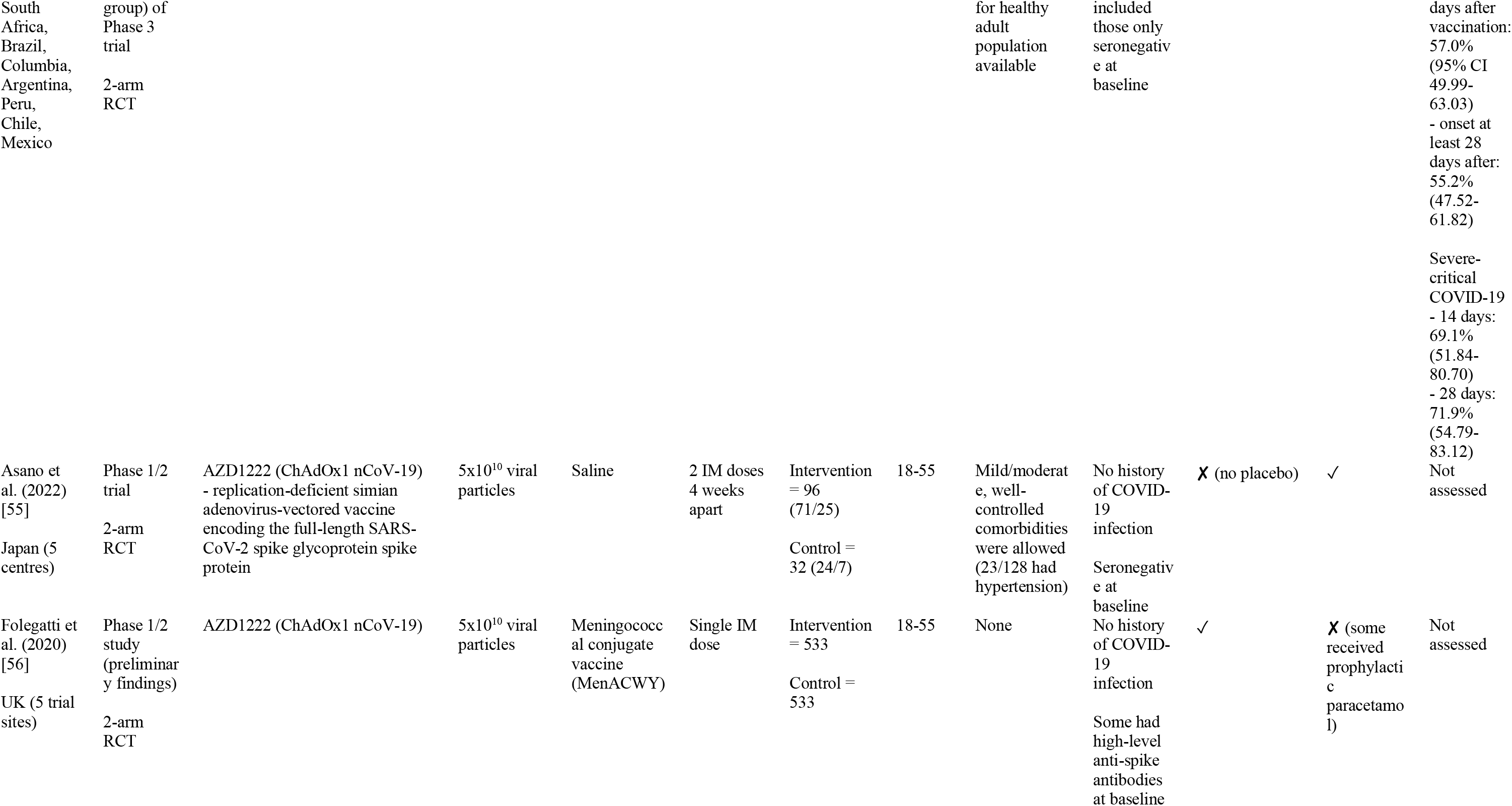
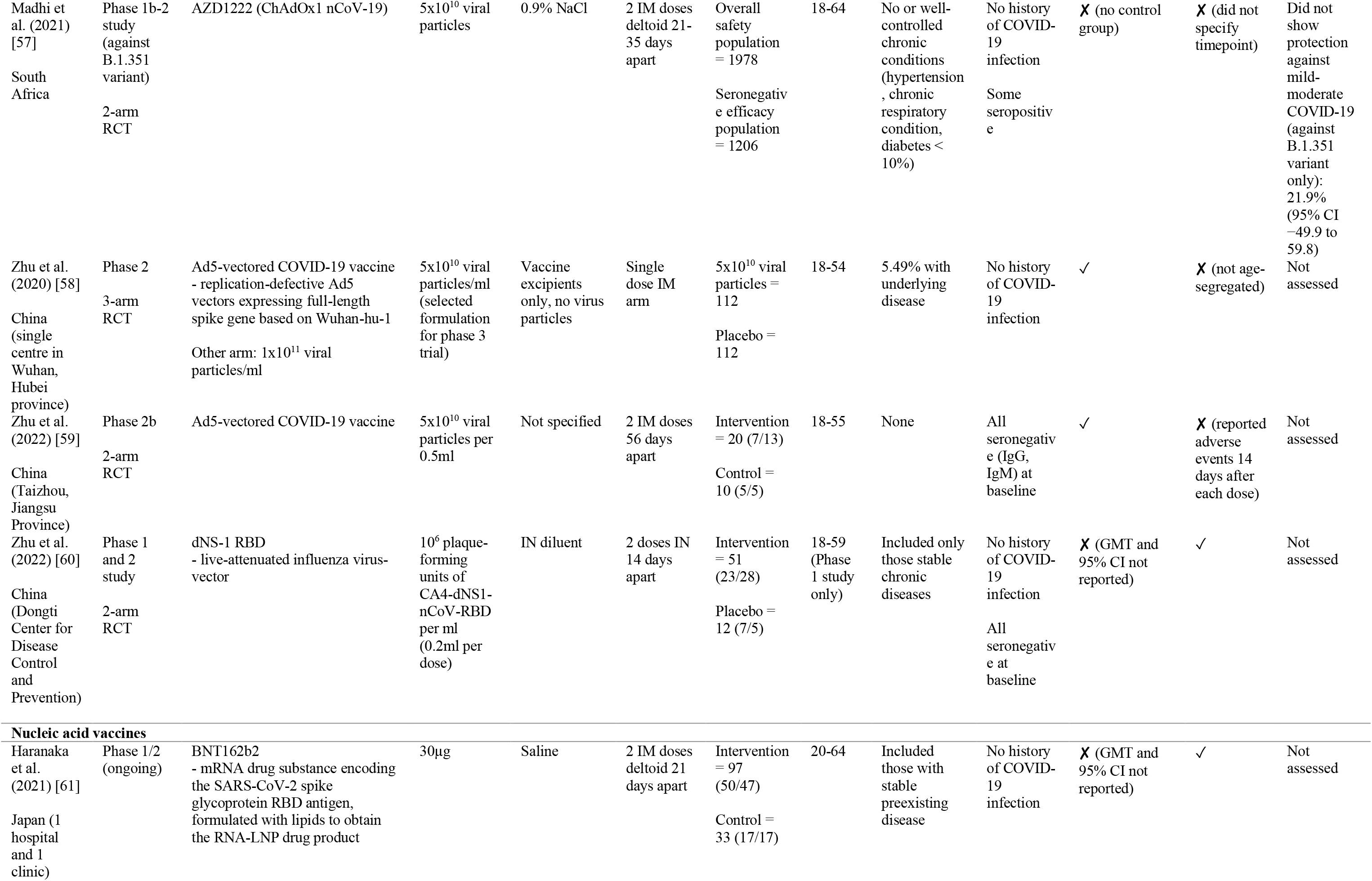
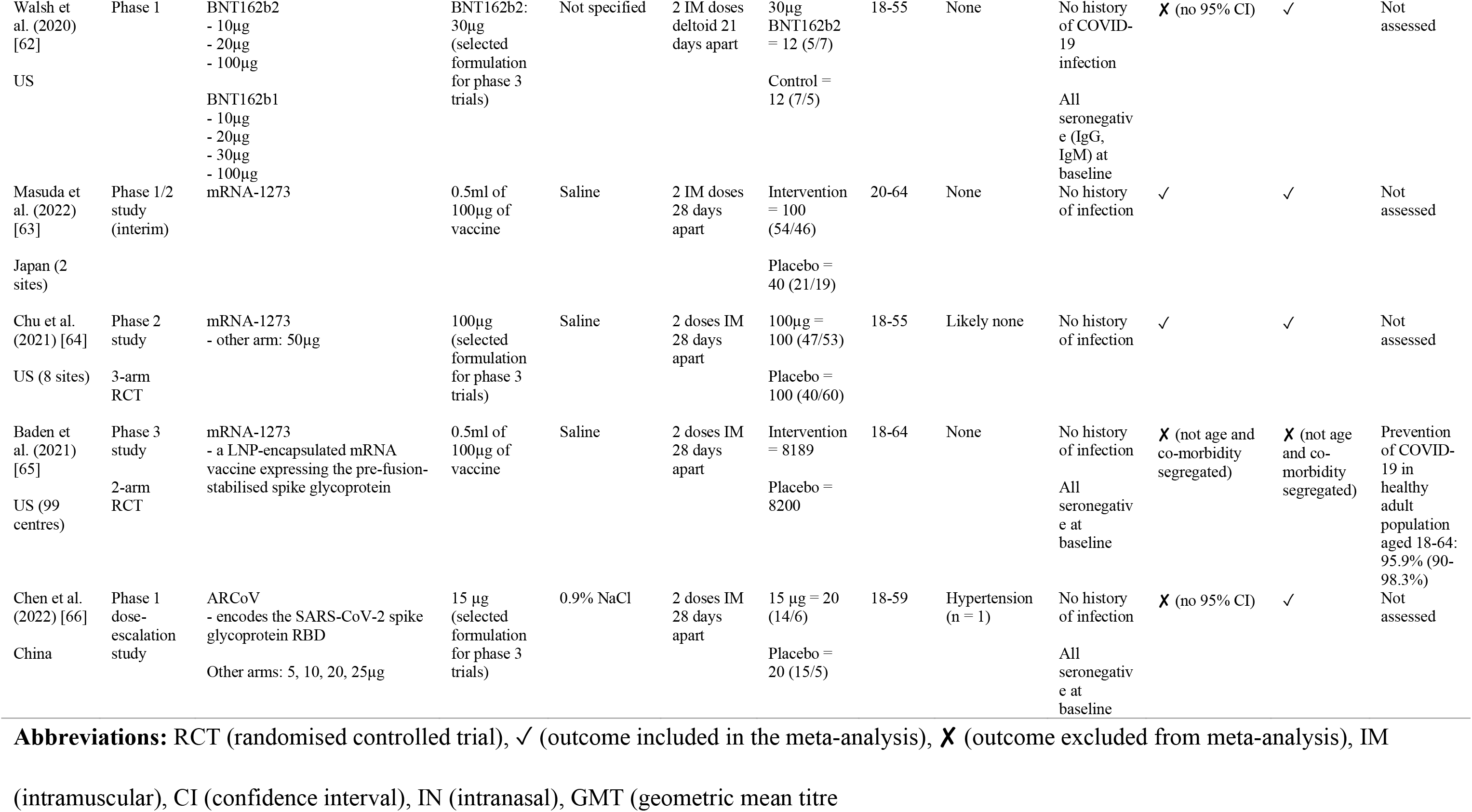
Characteristics of included studies.

### 3.3. Risk of bias

Most studies had some concerns (n = 31) with high ROB, while the rest had low ROB (n = 15). We rated the studies according to the RCT phase if possible; hence the total number does not add up to 41. Studies were rated with some concerns commonly due to the lack of information on allocation sequence concealment, and some studies did not specify the method of randomisation (Figure 2).

**Fig 2.**
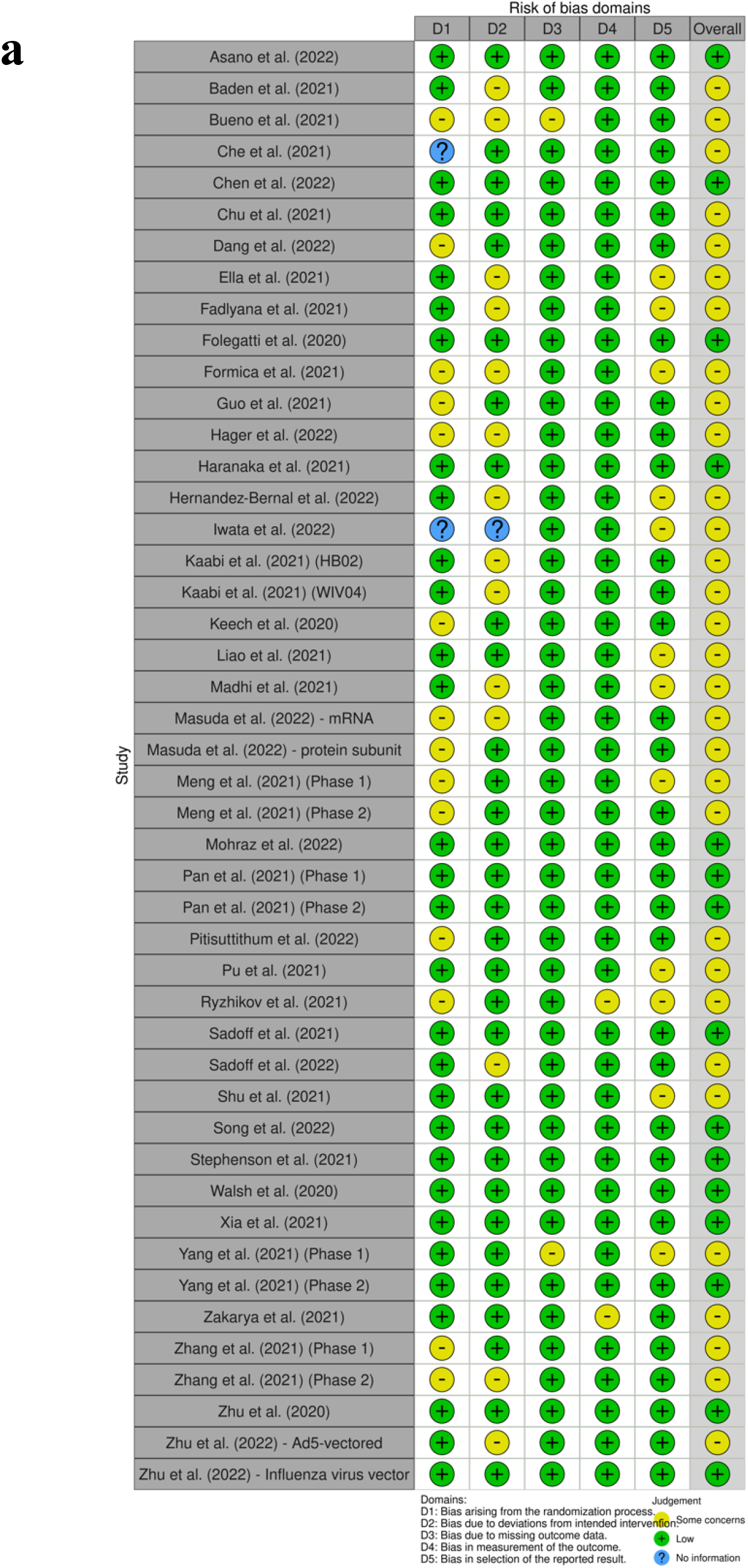
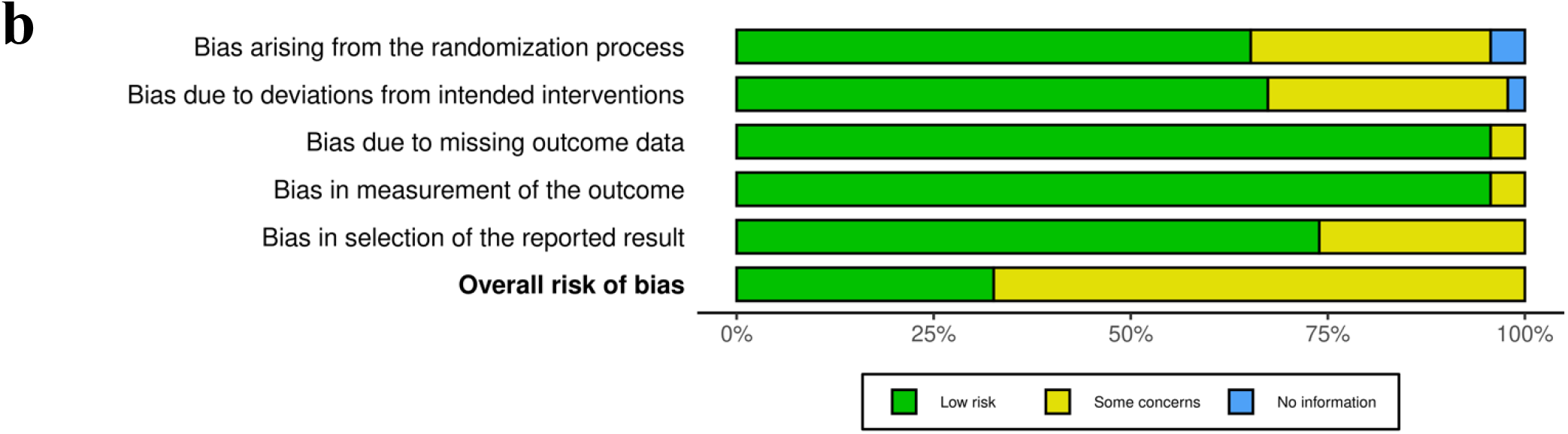
(a) Risk of bias rating for each study and (b) risk of bias rating for each domain across all studies.

### 3.4. GRADE assessment

Of the 16 outcomes assessed in the meta-analyses, 14 had moderate or high certainty of evidence. Certainty of evidence was downgraded most commonly due to high heterogeneity (inconsistency) and/or insignificant effect sizes (imprecision), and some outcomes were upgraded due to large effect sizes. The detailed GRADE assessment for each outcome is presented in Supplementary File 2.

### 3.5. Synthesis findings

Subgroup analysis was conducted based on vaccine type only, as age and sex were not possible due to inadequate information reported. Unless otherwise specified, sensitivity analysis confirmed the robustness of the results as the significance of the effect size remained unchanged.

#### 3.5.1. Immunogenicity outcomes

Cellular immune responses to COVID-19 vaccines are summarised in Table 2. All immunogenicity outcomes in the following meta-analyses refer to the number of days after the completion of the primary vaccine series (either two or three doses).

**Table 2.** Cellular immune responses of different COVID-19 vaccines.

| Vaccine | Author (year) | Assay methods | Findings |
| --- | --- | --- | --- |
| **Protein Subunit Vaccines** |  |  |  |
| **NVX-CoV2373** | Keech et al. (2020) [29] | Intracellular cytokine staining of antigen-specific CD4+ T cells | - Adjuvanted regimens induced antigen-specific polyfunctional CD4+ T-cell responses reflected in IFN-γ, IL-2 and TNF-α production on spike protein stimulation (strong bias towards Th1 phenotype)<br>- Th2 responses minimal (IL-5 and IL-13 cytokines) |
| **NDV-HXP-S** | Pitisuttithum et al. (2022) [33] | IFN-γ, IL-5 using ELI-Spot kit | IFN-γ/IL-5 ratio strongly skewed to Th1 response relative to the prevaccination baseline, suggesting the vaccine-induced T-cell memory capable of an antiviral response |
| **S-268019-b** | Iwata et al. (2022) [37] | IFN-γ, IL-2, IL-4, IL-5 in CD4+ or CD8+ cells using intracellular cytokine staining by flow cytometry, IFN-γ ELISpot assay | - Both vaccine regimens induced antigen-specific polyfunctional CD4+ T-cell responses reflected in IFN-γ, IL-2, IL-4 production on spike protein stimulation<br>- Strong bias towards Th1 phenotype<br>- Th2 responses minimal (IL-4 and IL-5)<br>- Substantial increase in IFN-γ levels observed on day 36 and 50 for those receiving vaccine |
| **Sf9 cells** | Meng et al. (2021) [38] | Enzyme-linked immunospot (ELISpot) | - In the phase 1 trial, the positive rate of IFN-γ peaked at 14 days than that at 28 days after the last dose vaccination.<br>- Among three vaccine groups, no significant difference was found in the positive rate of IFN-γ at 14 days after the last dose vaccination, with 50% in the adult low dose group (0, 28 days), 88% in the adult high dose group (0, 28 days), 88% in the adult high dose group (0, 14, 28 days), and 8% in the adult placebo group. |
| **GBP510 with/without AS03** | Song et al. (2022) [69] | Intracellular cytokine staining using SARS-CoV-2 RBD | - GBP510 adjuvanted with AS03 induced stronger CD4+ T-cell responses with higher percentages of IFN-γ, TNF-α, and IL-2 expression compared to unadjuvanted GBP510.<br>- IL-4 was inconsistent and IL-5 was nearly inexistent across all groups. |
| **Inactivated Vaccines** |  |  |  |
| **BBV152** | Ella et al. (2021) [43] | - ELISpot<br>- Intracellular cytokine staining | - IFN-γ ELISpot responses against SARS-CoV-2 peptides peaked at about 100–120 spot-forming cells per million peripheral blood mononuclear cells in all vaccinated groups on day 28.<br>- The Algel-IMDG groups elicited CD3+, CD4+, and CD8+ T-cell responses that were reflected in the IFN-γ production<br>- a minimal detection of less than 0.5% of CD3+, CD4+, and CD8+ T-cell responses in the 6 μg with Algel group and the Algel group (placebo). |
| **CoronaVac** | Bueno et al. (2021) [47] | ELISPOT and flow cytometry assays were performed using isolated peripheral blood mononuclear cells (PBMCs). | - Immunisation with CoronaVac induces a T-cell response polarised toward a Th1 immune profile, as the secretion of interleukin-4 by T cells was mainly undetected.<br>- Modest increases in the expression of activation-induced markers were detected for both MP-CD8A and MP-CD8B. |
| **inactivated SARS-CoV-2 vaccine** | Pu et al. (2021) [49] | - Human IFN-γ ELISpot Kit<br>- Bio-Plex Pro Human Cytokine 48-Plex | - The specific positive cytotoxic T lymphocyte (CTL) responses against the S protein, N protein and virion in the ELISpot assay indicated a distinct increase at day 28 after the booster for both schedules. These results suggest that the vaccine elicits a synchronous dynamic response involving antibodies and CTLs against the viral antigens.<br>- There were no significant differences between the vaccine and placebo groups with regard to the counts of various T cell populations in the peripheral blood. |

| **Virus Vector Vaccines** |  |  |  |
| --- | --- | --- | --- |
| **Ad26.COV2.S** | Sadoff et al. (2021) [52] | Intracellular cytokine staining with the use of two pools of S-peptide pools of 15 mers overlapping by 11 amino acids | - In cohort 1a, Th1 response to S peptides was detected in 76% (95% CI 65-86) of low-dose recipients and in 83% (95% CI, 73 to 91) of high-dose recipients; the corresponding values in cohort 3 were 60% (95% CI 46-74) and 67% (95% CI 53-79), respectively.<br>- In cohort 1a, the median CD4<sup>+</sup> Th1 response to S peptides increased from an undetectable level at baseline to a median of 0.08% (IQR 0.05-0.16) in low-dose recipients and 0.11% (IQR, 0.07-0.16) in high-dose recipients on day 15; in cohort 3, the corresponding values were 0.09% (IQR 0.04-0.17) and 0.11% (IQR 0.04-0.15), respectively.<br>- S-specific CD8<sup>+</sup> T-cell responses, as identified by the expression of interferon-γ or interleukin-2 cytokines on S-peptide stimulation. On day 15 in cohort 1a, CD8<sup>+</sup> T-cell response was detected in 51% of participants (95% CI 39-63) in the low-dose group and in 64% (95% CI 52-75) in the high-dose group, with a median S-specific CD8<sup>+</sup> T-cell response of 0.07% (IQR 0.03-0.19) and 0.09% (IQR, 0.05-0.19), respectively.<br>- In cohort 3, CD8<sup>+</sup> T-cell responses were lower, with an incidence of 36% (95% CI 23-51) in the low-dose group and 24% (95% CI 13-37) in the high-dose group, with a median response of 0.06% (IQR 0.02-0.12) and 0.02% (IQR 0.01-0.08), respectively. |
| **Ad26.COV2.S** | Stephenson et al. (2021) [53] | - IFN-γ and IL-4 ELISPOT assays<br>- Multiparameter ICS assays | - IFN-γ ELISPOT responses were observed in 65% (13/20) of vaccine recipients by day 15 and in 84% (16/19) of vaccine recipients by day 71, with no significant differences among groups<br>- No IL-4 responses were observed, indicating a TH1-biased cellular immune response.<br>- Multiparameter ICS assays confirmed the<br>induction of central memory<br>CD27+/CD45RA-/CD4+ and CD8+ T-cell responses. |
| **AZD1222 (ChAdOx1 nCoV-19)** | Folegatti et al. (2020) [56] | Ex-vivo interferon-γ ELISpot assay | - Interferon-γ ELISpot responses against SARS-CoV-2 spike peptides peaked at 856 spot-forming cells per million peripheral blood mononuclear cells (IQR 493–1802; n=43) at day 14, declining to 424 (221–799; n=43) by day 56 after vaccination. |
| **AZD1222 (ChAdOx1 nCoV-19)** | Madhi et al. (2021) [57] | ImmunoSEQ® Assay | - The ChAdOx1 nCoV-19 vaccine caused the expansion of CD4+ and CD8+ T lymphocytes to specific epitopes of the spike protein.<br>- D215G mutation found in the B.1.351 variant is within a region that had prevalent T-cell antigen responses |
| **Ad5-vectored COVID-19 vaccine** | Zhu et al. (2020) [58] | IFN-γ ELISpot | - Significant activation in postvaccination T-cell responses in terms of spot-forming cells observed both in participants with high and low pre-existing neutralising antibodies at day 28.<br>- Baseline ELISpot T-cell responses were negative in 506 (>99%) of 508 participants. Ad5-vectored COVID-19 vaccine induced significant SARS-CoV-2 spike glycoprotein-specific IFN-γ ELISpot responses in 227 (90%, 95% CI 85–93) of 253 participants receiving the $1 \times 10^{11}$ viral particles dose, and 113 (88%, 81–92) of 129 participants receiving the $5 \times 10^{10}$ viral particles dose at day 28.<br>- A median of 11.0 spot-forming cells (IQR 5.0–25.0) and 10.0 spot-forming cells (6.0–21.0) per $1 \times 10^5$ peripheral blood mononuclear cells in participants in the $1 \times 10^{11}$ viral particles and $5 \times 10^{10}$ viral particles dose groups, respectively, were observed at day 28, with increases of more than 10-times in both dose groups.<br>- The IFN-γ-ELISpot responses were not significantly different between the dose groups at day 28. - No positive IFN-γ ELISpot T-cell responses were detected in the placebo group postvaccination. |
| **dNS-1 RBD** | Zhu et al. (2022) [60] | INF- $\gamma$ ELISpot | - No specific T-cell responses were detected in PBMCs from vaccinators 1 month after the participants had received the second dose. |
| **Nucleic Acid Vaccines** |  |  |  |
| **ARCoV** | Chen et al. (2022) [66] | ELISpot | - Following the first vaccination, only a small proportion of participants in each vaccine group was positive for IFN-γ-expressing cells. However, after the second vaccination, all participants in the 5 μg, 10 μg, 15 μg, and 20 μg groups were positive for IFN-γ- expressing cells.<br>- All participants were positive with IL-2-expressing cells at day 7 after the second vaccination. |

##### 3.5.1.1. Neutralising antibodies (live virus neutralisation)

Four studies reported neutralising antibody levels at 7 days after vaccination (n = 281) [34, 38, 44, 67], which was significantly higher in the vaccinated group compared to the control group (SMD = 2.51, 95% CI 1.58-3.44, p < 0.00001). Heterogeneity was considerable (I² = 84%, p = 0.0004) (Figure 3a).

**Fig. 3.**
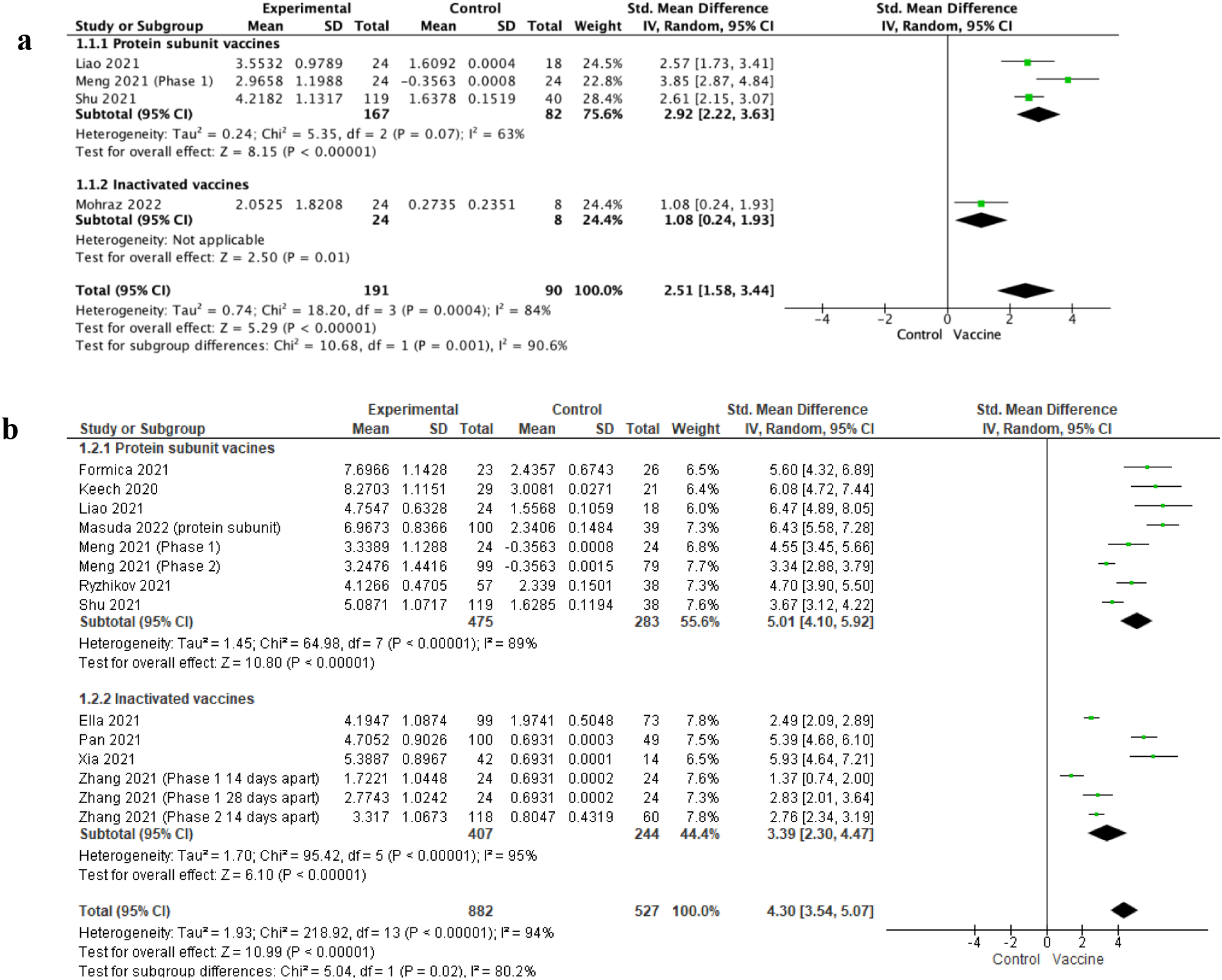
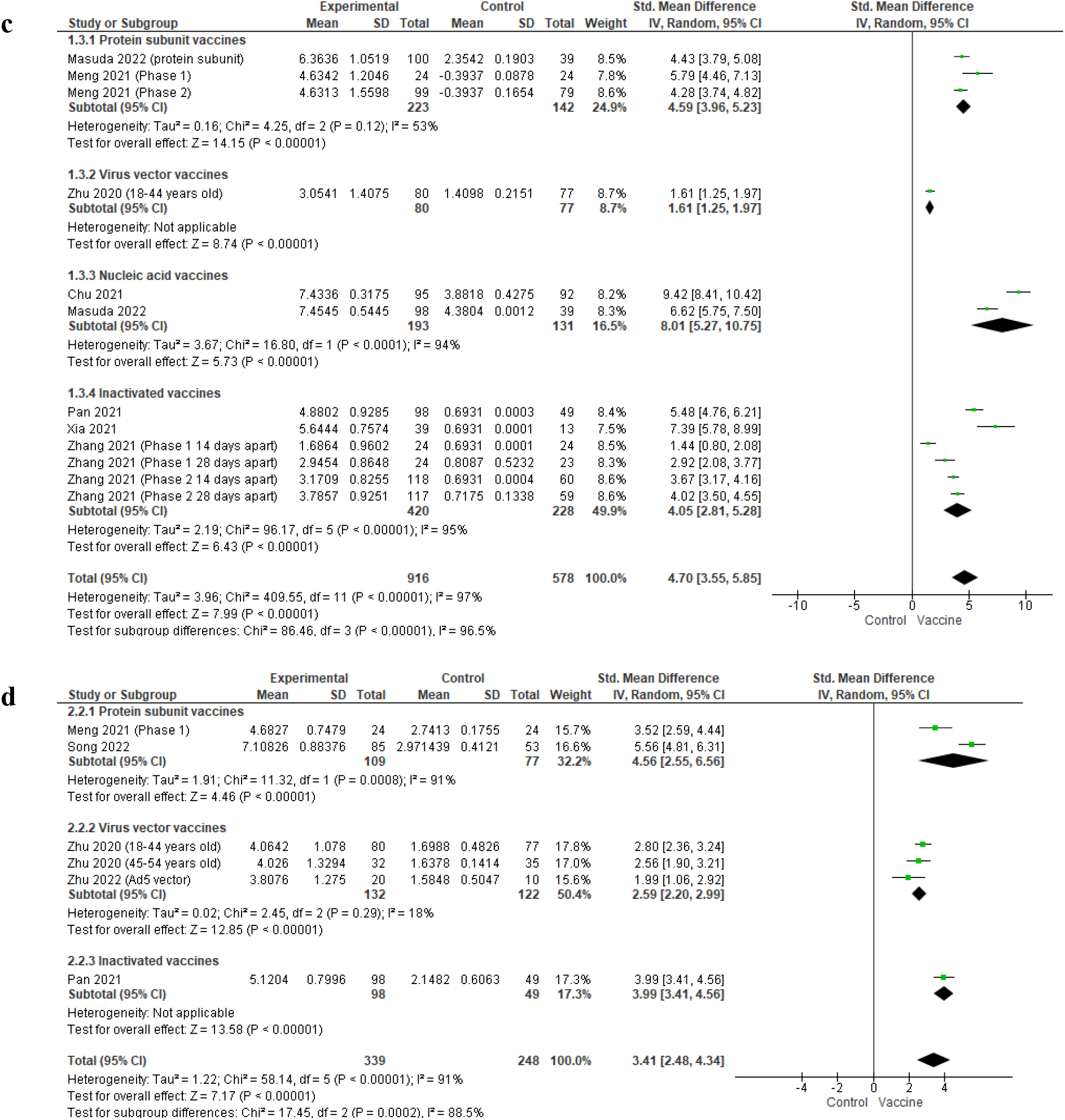
(a) Forest plot for meta-analysis of log-transformed neutralising antibody levels 7 days after COVID-19 vaccination (measured using live virus neutralisation assays). (b) Forest plot for meta-analysis of log-transformed neutralising antibody levels 14 days after COVID-19 vaccination (measured using live virus neutralisation assays). (c) Forest plot for meta-analysis of log-transformed neutralising antibody levels 28 days after COVID-19 vaccination (measured using live virus neutralisation assays). (d) Forest plot for meta-analysis of log-transformed neutralising antibody levels 28 days after COVID-19 vaccination (measured using pseudo-neutralising antibody assays)

At 14 days after vaccination (n = 1409, 11 studies) [29–31, 34, 38, 39, 43, 45, 46, 67, 68], neutralising antibodies were significantly higher in the vaccinated group than in the control group (SMD = 4.30, 95% CI 3.54-5.07, p < 0.00001). Heterogeneity was also considerable (I² = 94%, p < 0.00001), and there was a significant subgroup difference (I² = 80.2%, p = 0.02). Protein subunit vaccines induced higher levels of neutralising antibodies (SMD = 5.01, 95% CI 4.10-5.92, p < 0.00001) than inactivated vaccines (SMD = 3.39, 95% CI 2.30-4.47, p < 0.00001) (Figure 3b). Publication bias is likely as both Begg’s (p = 0.007) and Egger’s test (p < 0.001) were significant (Supplementary File 3 Figure S1)

At 28 days after vaccination (n = 1494, 8 studies) [30, 38, 45, 46, 58, 63, 64, 68], neutralising antibodies were significantly higher in the vaccinated group than in the control group (SMD = 4.70, 95% CI 3.55-5.85, p < 0.00001). Heterogeneity was considerable (I² = 97%, p < 0.00001) (Figure 3c).

##### 3.5.1.2. Neutralising antibodies (pseudovirus neutralisation)

Five studies reported neutralising antibodies at 28 days after vaccination [38, 45, 58, 59, 69], which was significantly higher in the vaccinated group than the control group (SMD = 3.41, 95% CI 2.48-4.34, p < 0.00001). Heterogeneity was considerable (I² = 91%, p < 0.00001) (Figure 3d).

##### 3.5.1.3. Anti-RBD IgG

Log-transformed anti-RBD IgG levels 14 days after vaccination (n = 1130, 8 studies) [34, 43–46, 58, 67, 69] were also significantly higher in the vaccinated group compared to the control group (SMD = 5.68, 95% CI 3.95-7.42, p < 0.00001) with considerable heterogeneity (I² = 99%, p < 0.00001) (Figure 4a).

**Fig. 4.**
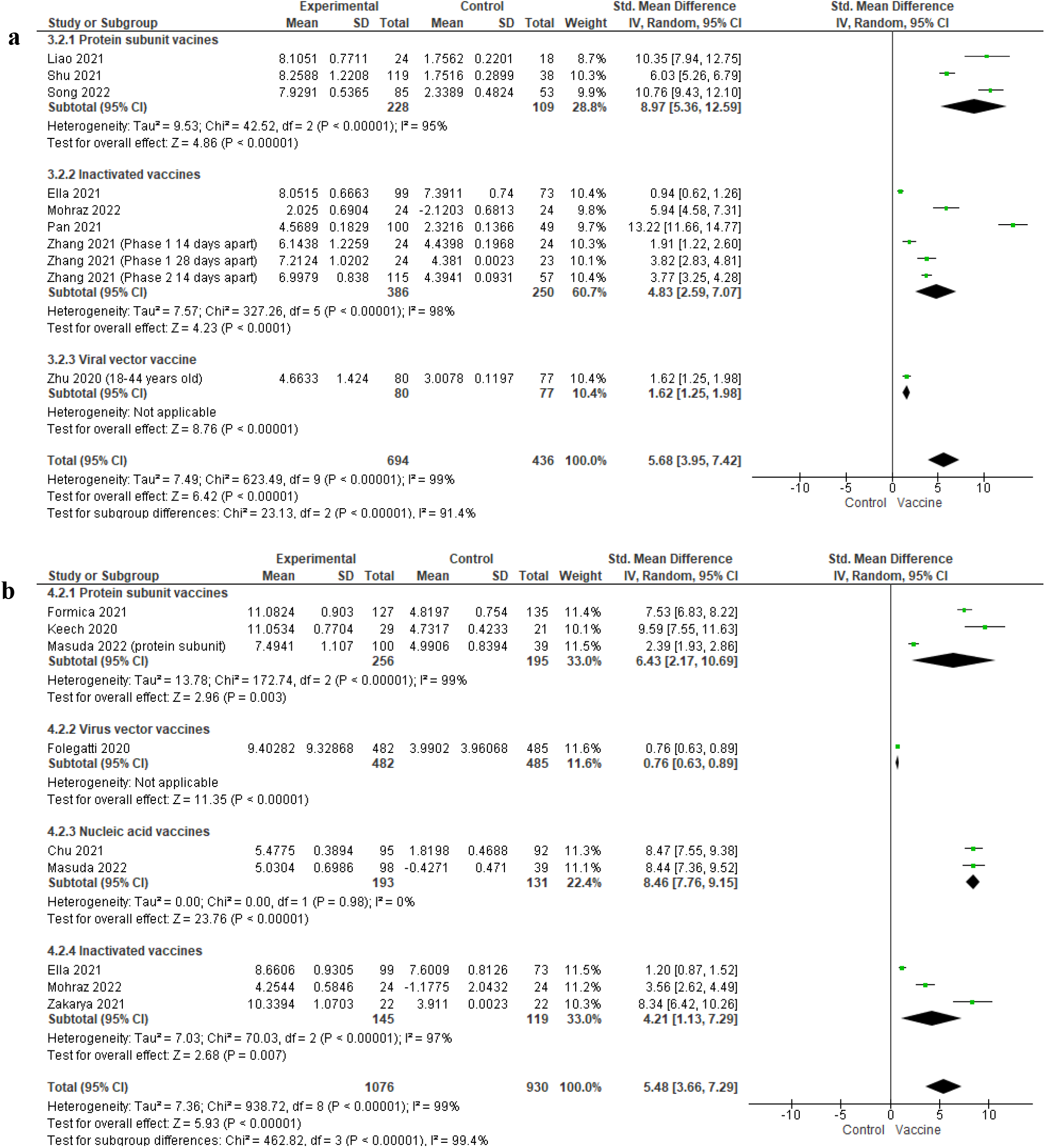
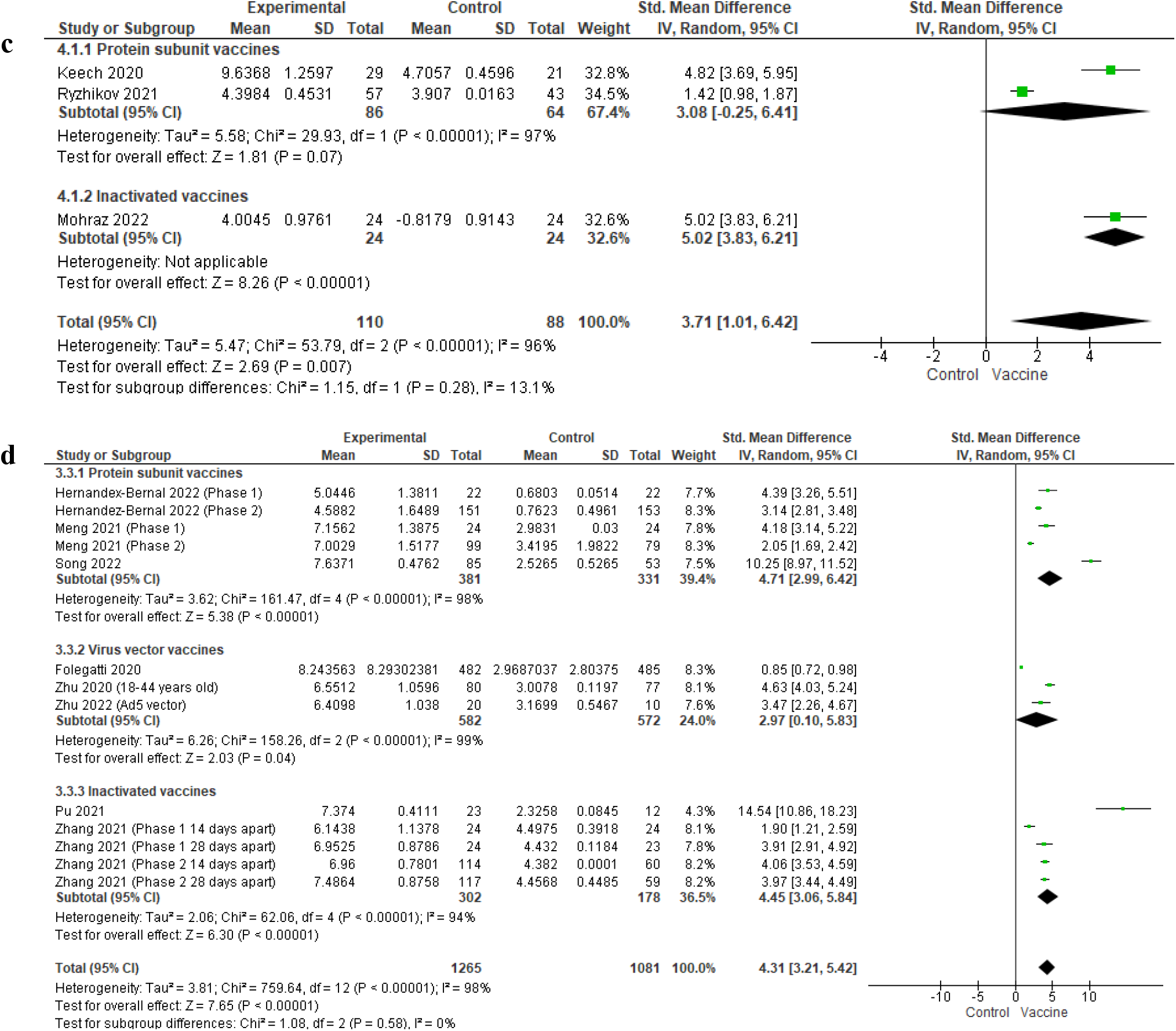
(a) Forest plot for meta-analysis of log-transformed anti-RBD IgG levels 14 days after COVID-19 vaccination. (b) Forest plot for meta-analysis of log-transformed anti-S IgG levels 14 days after COVID-19 vaccination. (c) Forest plot for meta-analysis of log-transformed anti-S IgG levels 7 days after COVID-19 vaccination. (d) Forest plot for meta-analysis of log-transformed anti-RBD IgG levels 28 days after COVID-19 vaccination

Log-transformed anti-RBD IgG levels 28 days after vaccination (n = 2326, 8 studies) [36, 38, 46, 49, 56, 58, 59, 69] was also significantly higher in the vaccinated group compared to the control group (SMD = 4.31, 95% CI 3.21-5.42, p < 0.00001). Heterogeneity was considerable (I² = 98%, p < 0.00001) (Figure 4b).

##### 3.5.1.4. Anti-S IgG

Three studies reported anti-S IgG levels at 7 days after vaccination (n = 198) [29, 39, 44], and anti-S IgG levels were significantly higher in the vaccinated group than the control group (SMD = 3.71, 95% CI 1.01-6.42, p = 0.007) with considerable heterogeneity (I² = 96%, p < 0.00001) (Figure 4c).

At 14 days after vaccination (n = 2006, 9 studies) [29–31, 43, 44, 51, 56, 63, 64], pooled SMD for anti-S IgG levels was 5.48 (95% CI 3.66-7.29, p < 0.00001) with considerable heterogeneity (I² = 99%, p < 0.00001) (Figure 4d).

#### 3.5.2 Safety outcomes

##### 3.5.2.1. Seven days after the first dose

Twelve studies reporting local adverse events seven days after the first dose of a COVID-19 vaccine were included in the meta-analysis (n = 1301) [31–33, 37, 39, 43, 44, 49, 51, 55, 64, 67], and those in the vaccine arm had a significantly higher risk of local adverse events compared to the control (pooled RR = 2.88, 95% CI 1.78-4.67, p < 0.0001). Heterogeneity was substantial (I² = 71%, p < 0.00001). There was a significant subgroup difference based on vaccine type (p = 0.03, I² = 65.7%), and only the inactivated vaccines subgroup showed an insignificant pooled RR of 1.43 (95% CI 0.60-3.41, p = 0.42), indicating that risk of local adverse events was similar between vaccine and control groups (Figure 5a). Publication bias is unlikely as Egger’s regression (p = 0.471) and Begg’s test (p = 0.638) were insignificant (Supplementary File 3 Figure S2). When the article by Mohraz et al. [44] was excluded during sensitivity analysis, heterogeneity became insignificant (I² = 17%, p = 0.28).

**Fig. 5.**
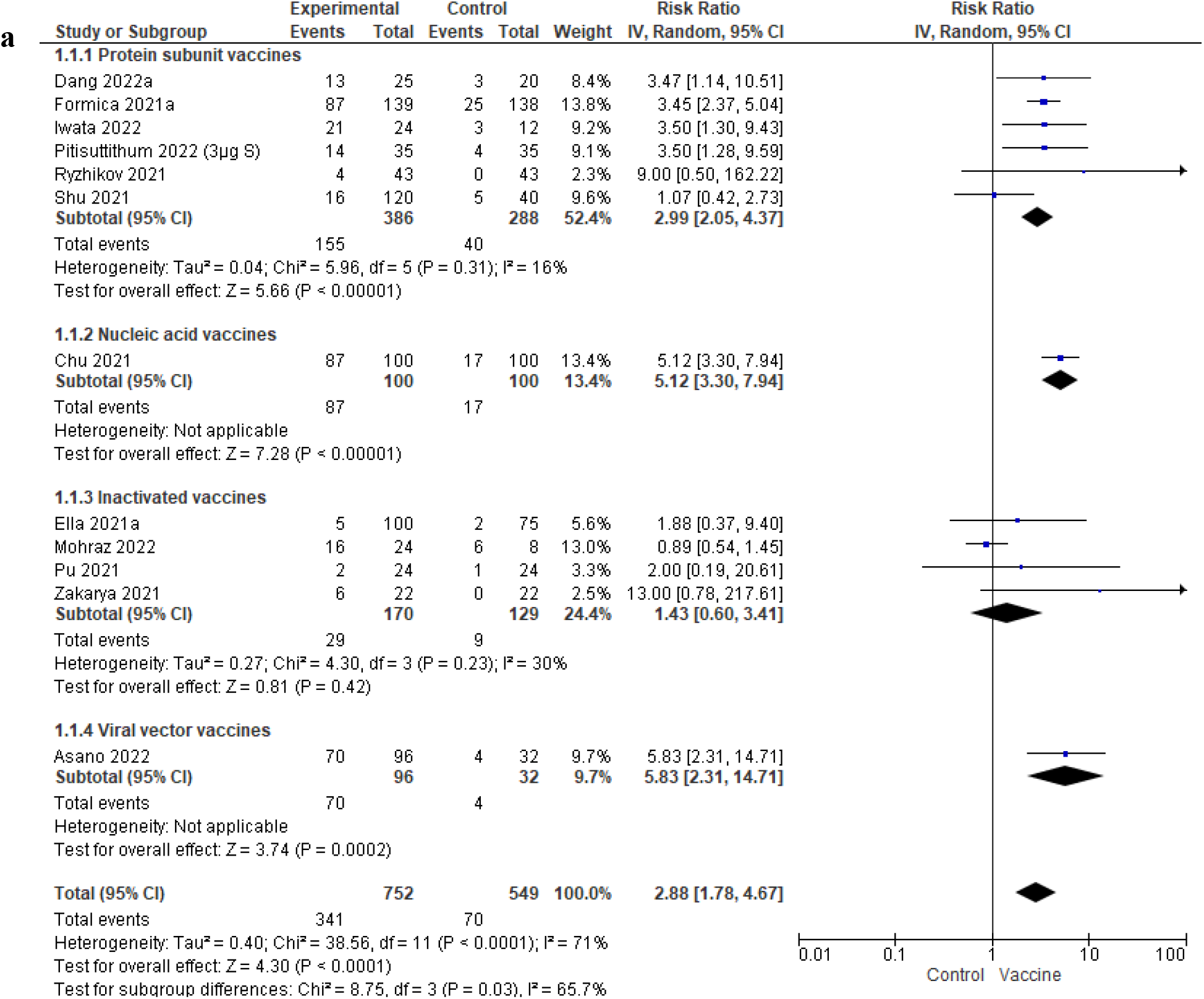
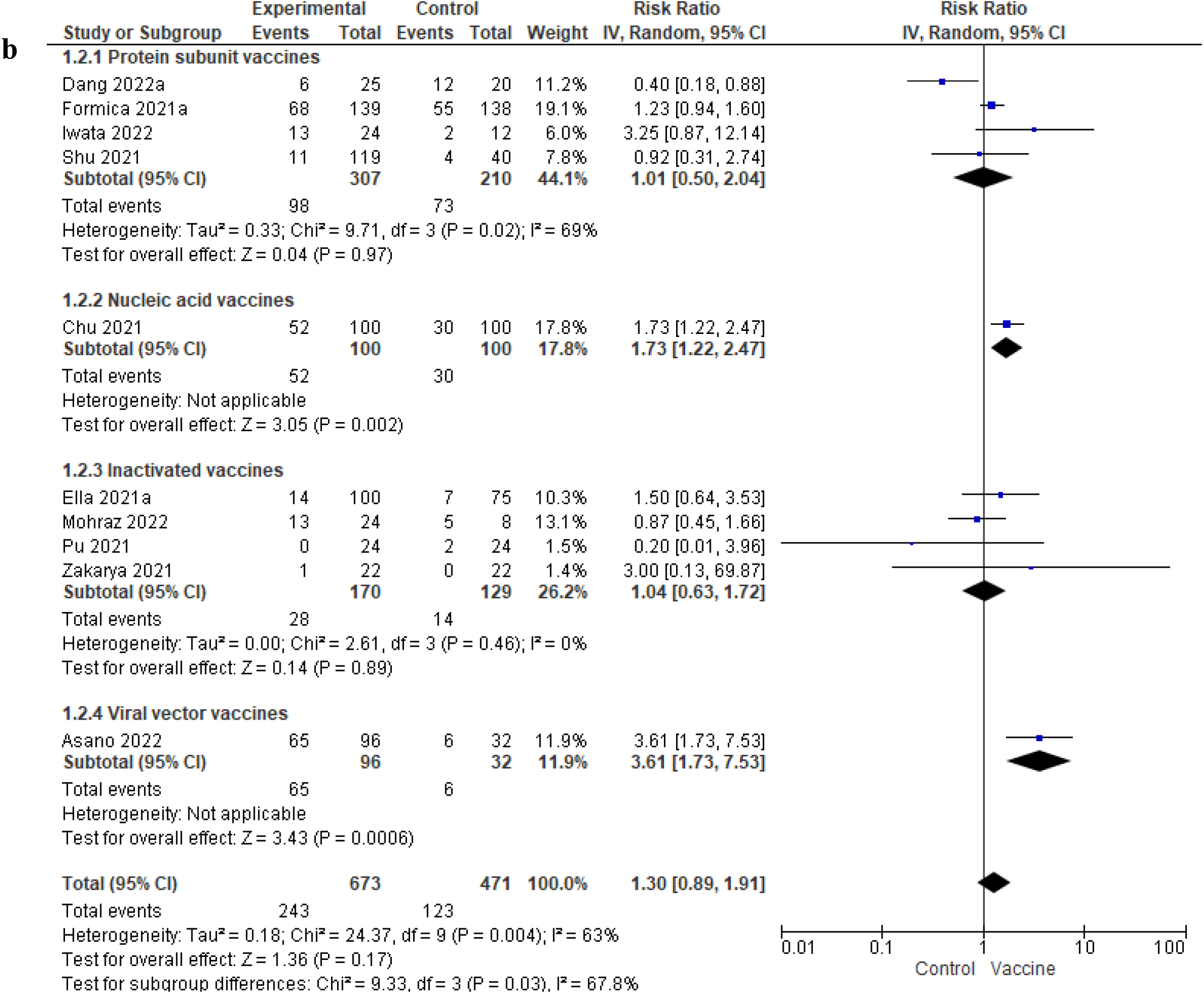
(a) Forest plot for meta-analysis of local adverse events after 7 days of the first COVID-19 vaccine dose. (b) Forest plot for meta-analysis of systemic adverse events after 7 days of first COVID-19 vaccine dose.

Ten studies reporting systemic adverse events seven days after the first dose of a COVID-19 vaccine were pooled (n = 1144) [31, 32, 37, 43, 44, 49, 51, 55, 64, 67], and the risk of systemic adverse events was similar between vaccine and control groups (pooled RR = 1.30, 95% CI 0.89-1.91, p = 0.17). Heterogeneity was substantial (I² = 63%, p = 0.004). There was also a significant subgroup difference based on vaccine type (p = 0.03, I² = 67.8%) (Figure 5b). Publication bias is unlikely as Egger’s regression (p = 0.452) and Begg’s test (p = 0.484) were insignificant (Supplementary File 3 Figure S3).

##### 3.5.2.1. Seven days after the second dose

Ten studies reporting local adverse events seven days after the second dose of a COVID-19 vaccine were pooled (n = 1193) [31–33, 37, 39, 43, 44, 55, 64, 67]. Similarly, RR was higher in the vaccine group (pooled RR = 2.61, 95% CI 1.38-4.90, p = 0.003), and heterogeneity is considerable (I² = 80%, p < 0.00001). A significant subgroup difference was found (p = 0.0003, I² = 84.2%), with only inactivated vaccines reporting an insignificant effect size (pooled RR = 1.05, 95% CI 0.48-2.28, p = 0.90) (Figure 6a). Publication bias is unlikely as Egger’s regression (p = 0.608) and Begg’s test (p = 0.862) were insignificant (Supplementary File 3 Figure S4).

**Fig. 6.**
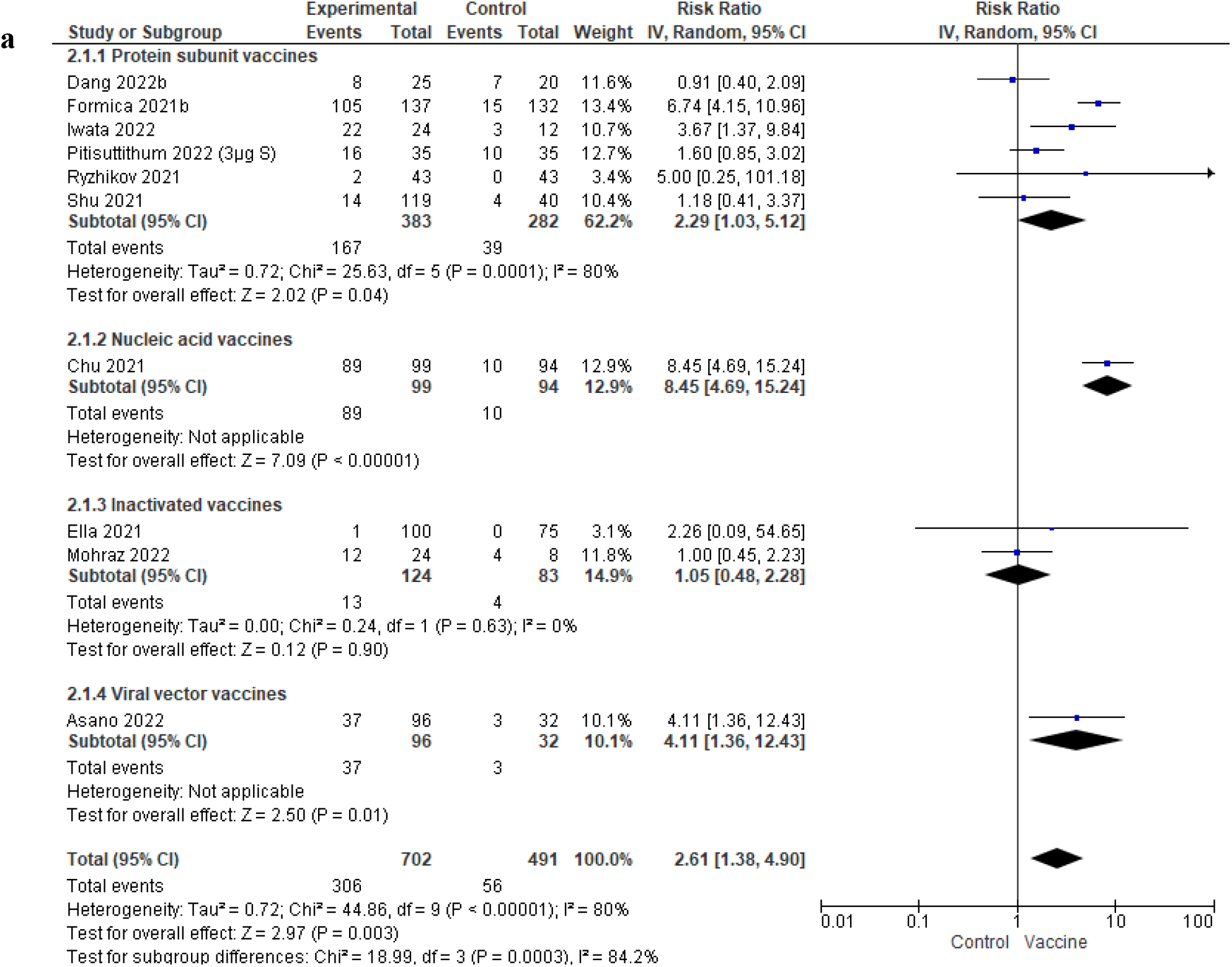
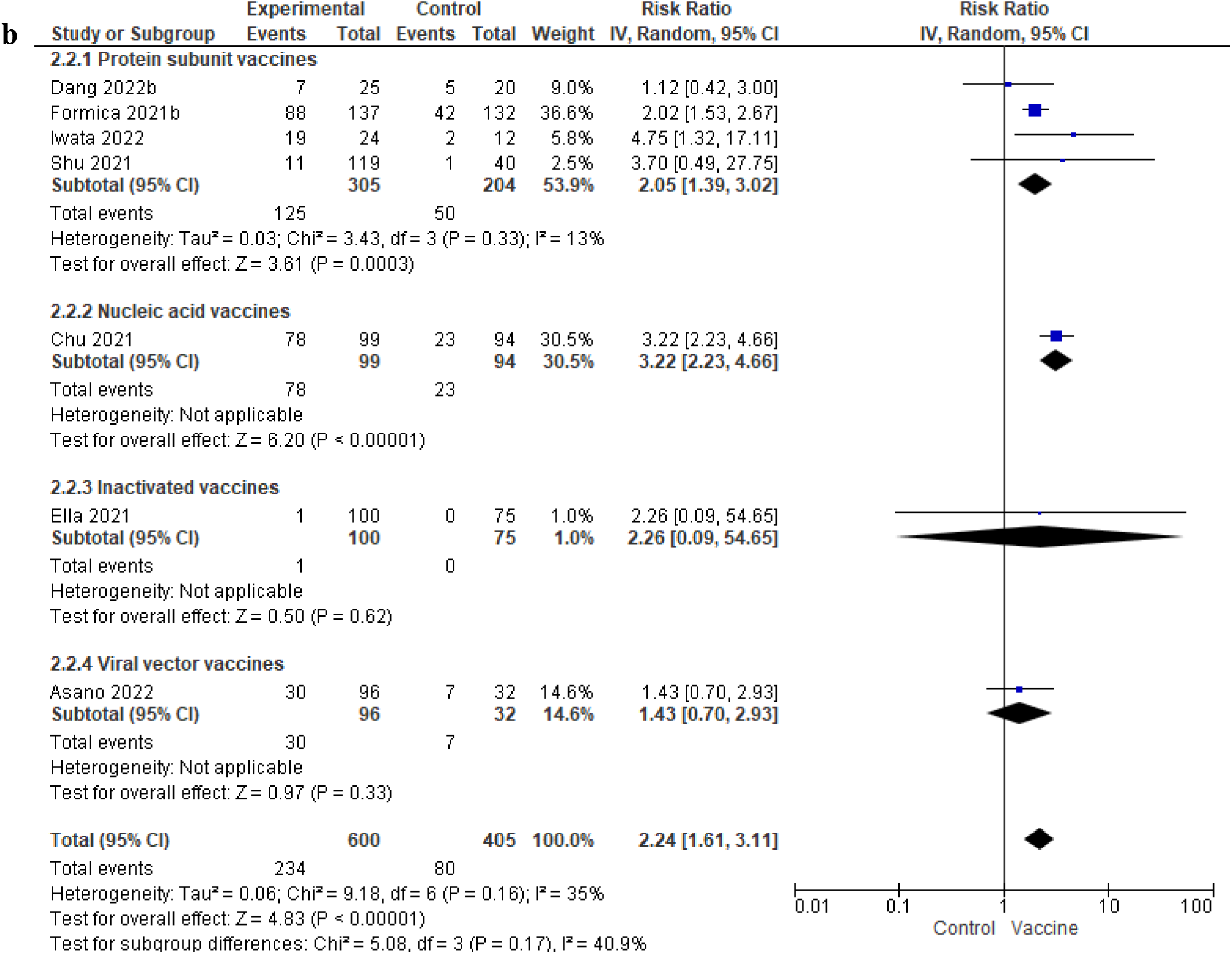
(a) Forest plot for meta-analysis of local adverse events after 7 days of the second COVID-19 vaccine dose. (b) Forest plot for meta-analysis of systemic adverse events after 7 days of the second COVID-19 vaccine dose.

Seven studies reported systemic adverse events seven days after the second dose of a COVID-19 vaccine (n = 1005) [31, 32, 37, 43, 55, 64, 67], and the risk ratio was higher in the vaccinated group (pooled RR = 2.24, 95% CI 1.61-3.11, p < 0.00001). Heterogeneity was insignificant (I² = 35%, p = 0.16) (Figure 6b).

##### 3.5.2.2. One month after the first dose

Six studies reporting any adverse events 1 month after the first dose were pooled (n = 397) [29, 32–34, 44, 67], and there were no significant differences between groups receiving vaccine or control (pooled RR = 1.04, 95% CI 0.66-1.65, p = 0.87). Heterogeneity was moderate (I² = 48%, p = 0.09) (Figure 7a).

**Fig. 7.**
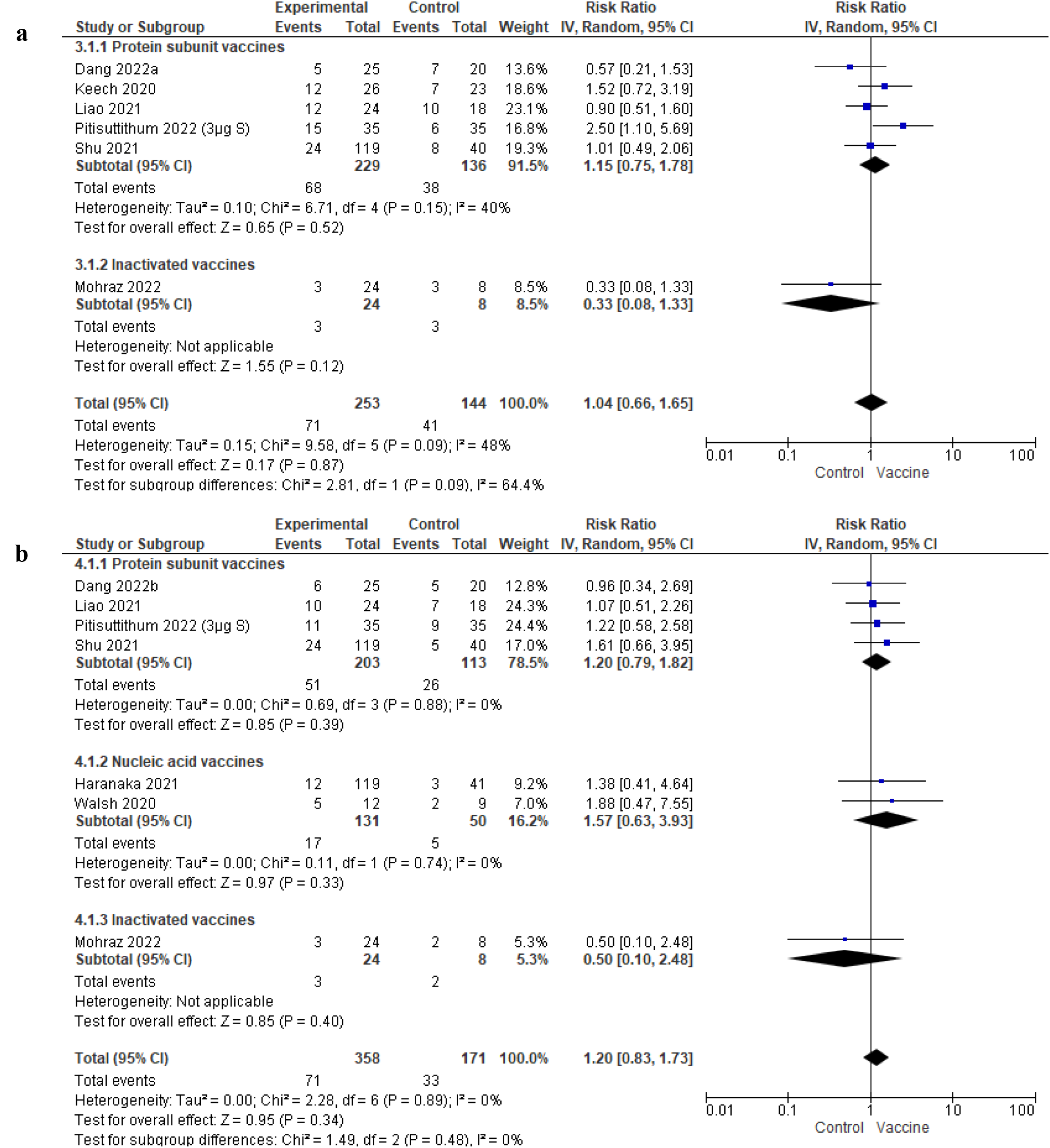
(a) Forest plot for meta-analysis of any adverse events after 1 month of the first COVID-19 vaccine dose. (b) Forest plot for meta-analysis of any adverse events after 1 month of the second COVID-19 vaccine dose.

##### 3.5.2.3. One month after the second dose

Seven studies reporting any adverse events 1 month after the second dose were pooled (n = 529) [32–34, 44, 61, 62, 67], and there were also no significant differences between the groups receiving vaccine or control (pooled RR = 1.20, 95% CI 0.63-1.73, p = 0.34). Heterogeneity was insignificant (I² = 0%, p = 0.89) (Figure 7b).

##### 3.5.2.4. Overall adverse events

Eight studies reported overall adverse events after 7 days (n = 1603) [38, 40, 41, 50, 52, 63, 66, 68], and the risk of adverse events was significantly higher in the vaccinated group (pooled RR = 1.68, 95% CI 1.21-2.34, p = 0.002). Heterogeneity was substantial (I² = 70%, p = 0.0001) (Figure 8a).

**Fig. 8.**
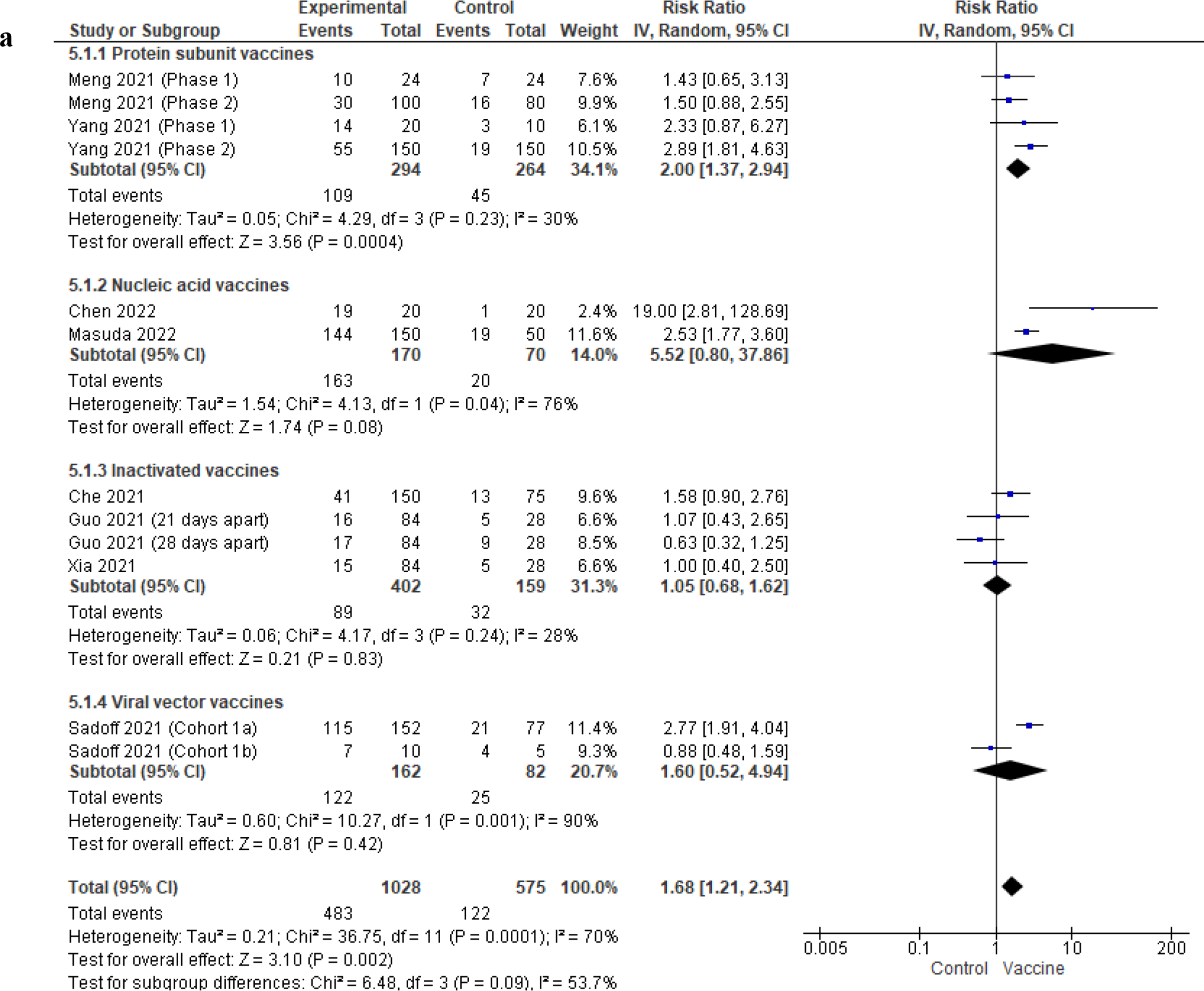
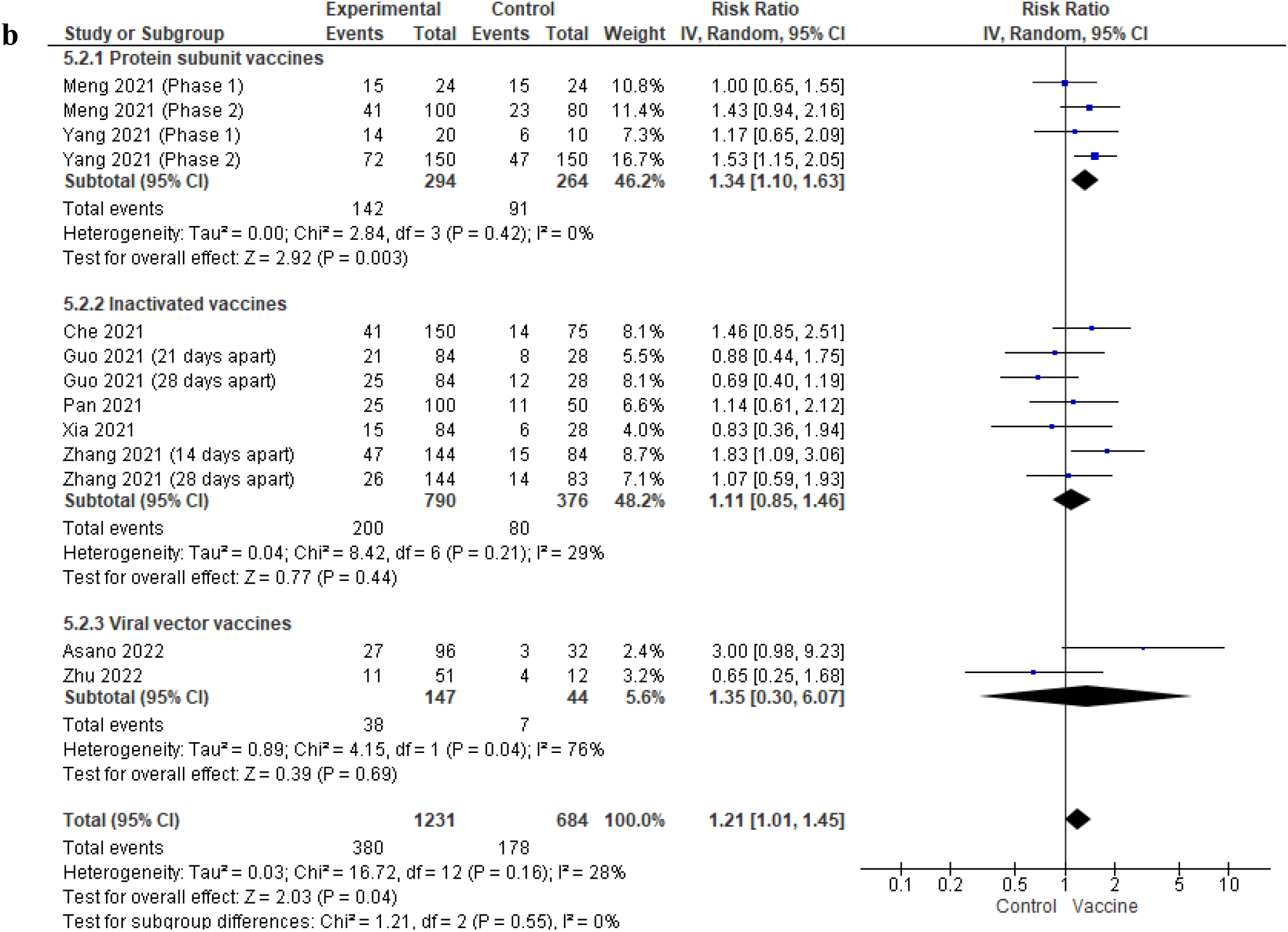
(a) Forest plot for meta-analysis for overall adverse events within 7 days post COVID-19 vaccination. (b) Forest plot for meta-analysis for overall adverse events within 1 month post COVID-19 vaccination

Nine studies reported overall adverse events after 1 month (n = 2235) [38, 40, 41, 45, 46, 50, 55, 59, 68], and the risk of adverse events was significantly higher in the vaccinated group (pooled RR = 1.19, 95% CI 1.01-1.40, p = 0.04). Heterogeneity was insignificant (I² = 26%, p = 0.17) (Figure 8b).

##### 3.5.2.5 Serious adverse events

Serious adverse events, defined as Grade 3 or worse, were reported in 19 studies [29, 31, 35, 40, 42, 44, 48, 52, 54–57, 59, 60, 63–67]. However, they were rare, and many studies did not specify if these were related to the vaccine. Nonetheless, the studies concluded that the vaccines had an acceptable safety profile.

#### 3.5.3. Efficacy outcomes

Efficacy outcomes were summarised in Table 1, and 6 studies reported efficacy outcomes [35, 42, 47, 54, 57, 65] ranging from 21.9% (95% CI −49.9 to 59.8) against mild-moderate COVID-19 [57] to 95.9% in preventing COVID-19 [65]. However, they were based on previous circulating variants of concern; hence the findings would not be representative of its efficacy in the current COVID-19 situation in which Omicron is the predominant strain, with subvariants such as BA.4 and BA.5 making up most of the world’s COVID-19 cases [70].

## 4. Discussions

This systematic review and meta-analysis found that the vaccinated individuals had significantly immunogenic to COVID-19 compared to the placebo. Although our meta-analyses confirmed that vaccines induce significantly higher immune responses compared to placebo up to 28 days after completion of the primary vaccination series, this does not necessarily correlate with better disease outcomes [6]. Efficacy outcomes in healthy adults, which were rarely reported in this review, should still be relied upon to assess the clinical utility of a vaccine.

COVID-19 vaccines in healthy adults, as assessed in this review, were relatively safe with minimal serious adverse events, which is consistent with previous large-scale observational studies and reviews [71–74] . Subgroup analyses suggest that inactivated vaccines may result in the lowest risk of adverse events among the four major vaccine genres. Similar incidences of adverse events concur with other observational studies as well [75, 76]. Due to misinformation, there is significant vaccine hesitancy worldwide. This review provides empirical evidence that vaccines are usually safe, countering the misconception-led vaccine hesitancy [77].

All meta-analyses conducted in this review found that immune responses (neutralising antibodies, anti-RBD and anti-S IgG) were significantly higher than the placebo group after vaccination. However, these measures may not all contribute to establishing immunity to COVID-19 infection and reducing the severity of COVID-19 disease [11]. Nonetheless, neutralising antibody levels are predictive of their protective efficacy, and we found that neutralising antibody levels were the highest in the nucleic acid vaccines subgroup (Figure 4D), which correlates to their high efficacy in preventing COVID-19 infection, as established by previous studies [11, 78]. In the context of the current COVID-19 pandemic, it was found that neutralising antibody levels were reduced by at least 1/10^th^ against the Omicron variant compared to the original strain [11]. Hence, the findings of this outcome should be interpreted with caution as most included studies were conducted when previous strains, such as the Alpha, Beta and Delta strains were more prevalent [79]. Immune responses and actual protection against COVID-19 infection and severe disease would thus be lower in real-world conditions. A large-scale observational study found that homologous primary vaccination with 2 doses of ChAdOx1 nCoV-19, BNT162b2 or mRNA-1273 resulted in vaccine effectiveness of 48.9% (95% CI 39.2 to 57.1), 65.5% (95% CI 63.9-67.0) and 75.1% (95% CI 70.8 to 78.7) respectively at 2-4 weeks against symptomatic disease against the Omicron variant [80].

We have also descriptively summarised the cellular immune responses of COVID-19 vaccines in Table 2, which shows that they predominantly induce a Th1-mediated immune response. Studies included in the review utilised a variety of assays and outcomes; hence meta-analysis was not possible. A recent study performing head-to-head comparisons of the immune responses of those receiving mRNA-1273, BNT162b2, Ad26.COV2.S or NVX-CoV2373 vaccines found that while antibody titers declined over 6 months, memory T cells and B cells were comparatively stable, suggesting that immune memory from vaccination remains intact [81]. T-cell responses also remain robust against the Omicron variant [82], suggesting that while vaccinations may be less effective in preventing infection due to less neutralising antibodies generated against emerging variants, they are still paramount in reducing disease severity through SARS-CoV-2 specific T cells facilitating early recognition of COVID-19 virus and mediating antiviral responses [83]. Recent COVID-19 vaccine research has thus focused on the effectiveness of heterologous and homologous boosters to make up for the natural decay of antibody levels over time [84].

To the best of our knowledge, this is the first systematic review and meta-analysis that focuses on the effects of COVID-19 vaccines in the healthy adult population and provides comprehensive evidence that current vaccines are safe and immunogenic in the healthy adult population, unlike early meta-analyses on COVID-19 vaccines which had pooled outcomes without accounting for the differences in participant characteristics between studies. In addition, we have included the most recent RCTs which were not included in the latest meta-analyses published [85, 86].

However, our review was not devoid of limitations. First, numerous studies could not be included in the review or be pooled in the meta-analysis as they did not provide subgroup analyses of the RCT results on the healthy adult population. Our meta-analyses thus had relatively small sample sizes, with most included studies being Phase 1 or 2 trials where sample sizes are smaller. Our subgroup analyses should be interpreted with caution as there was an uneven distribution of studies in each subgroup [28]. Second, we only included English language studies and could have missed out on studies in other languages. Third, due to the varied outcomes investigated and poor reporting of information by some studies, some findings could not be included in the meta-analyses (e.g., no 95% CI reported, different timepoints for outcome measurement).

## 5. Conclusions

Overall, this systematic review and meta-analysis show that COVID-19 vaccines are safe and immunogenic in the healthy adult population. Future individual patient data-driven meta-analyses should be conducted to fully utilise the available RCT data and provide a more comprehensive analysis of the effects of COVID-19 vaccines according to different patient characteristics (e.g., gender, ethnicities, comorbidities). Thorough longitudinal designs calibrating exposure to SARS-CoV-2 vaccine-mediated adaptive immunity in relation to the consequential long-term advantageous and detrimental impact on diverse ethnic populations can assist in refurbishing preemptive policies against the future occurrence and outbreak of COVID-19.

## Supporting information

Supplementary File 1. Search strategy

Supplementary File 2. GRADE evidence profile

Supplementary File 3. Supplementary meta-analysis figures

## Data Availability

This is meta-analysis and the collated dataset analysed in the present article are available upon reasonable request to the authors.

## Acknowledgements

We would like to thank Ms Zeng Bentuo for her assistance in searching for relevant articles and Dr Liliya Eugenevna Ziganshina for her valuable advice regarding the scoping of this review.

## Declarations

### Ethics approval

This is a systematic review and meta-analysis using publicly available data from peer-reviewed articles reposited in bibliographic databases; hence, no ethical approval is needed.

### Author contributions

All authors attest that they meet the International Committee of Medical Journal Editors (ICMJE) criteria for authorship. Si Qi Yoong: Methodology, Formal analysis, Investigation, Writing – original draft, Writing – review & editing, Visualisation. Priyanka Bhowmik: Conceptualisation, Investigation, Writing – review & editing. Debprasad Dutta: Conceptualisation, Writing – review & editing, Supervision.

