## Supplementary File 1. Search strategy for "Comparative Immunogenicity, Safety and Efficacy Profiles of four COVID-19 Vaccine types in healthy adults: Systematic Review cum Meta-analysis of Clinical Trial data"

### Comparative Immunogenicity, Safety and Efficacy profiles of four COVID-19 Vaccine types in healthy adults: Systematic Review cum Meta-analysis of Clinical Trial data

Si Qi YOONG[a], Priyanka BHOWMIK[b], Debprasad DUTTA[c,d,] *

**Authors Affiliations:**

Si Qi YOONG
[a] Alice Lee Centre for Nursing Studies, Yong Yoo Lin School of Medicine
National University of Singapore, Singapore

Priyanka BHOWMIK
[b] Tbilisi State Medical University, Georgia

Debprasad DUTTA*
[c] Mazumdar Shaw Center for Translational Research (MSCTR), Mazumdar Shaw Medical Foundation, Bangalore, India
[d] Institute of Infection, Veterinary and Ecological Sciences (IVES), University of Liverpool, Liverpool, UK


***Table S1. PICO Concept (PubMed Syntax and Logic)***

| **PICO** | **Concept** | **Search terms** | |
|---|---|---|---|
| | | *Index terms* | *Keywords* |
| Population | Adults | "Adult"[Mesh] OR "Young Adult"[Mesh] | "adult*"[Title/Abstract] OR "young adult"[Title/Abstract] |
| | | "Middle Aged"[Mesh] | "middle age*"[Title/Abstract] OR "age"[Title/Abstract] OR "older adult*"[Title/Abstract] |
| Intervention | COVID-19 vaccines | "COVID-19 Vaccines"[Mesh] | "covid 19 vaccine*"[Title/Abstract] OR "coronavirus vaccine*"[Title/Abstract] OR "sars cov 2 vaccine*"[Title/Abstract] OR "ncov vaccine*"[Title/Abstract] |
| Outcome | Immunogenicity, Safety, efficacy and effectiveness | "Immunity"[MeSH Terms] OR "immunity, cellular"[MeSH Terms] OR "immunity, heterologous"[MeSH Terms] OR "immunity, humoral"[MeSH Terms] | "immunit*"[Title/Abstract] OR "humoral immunit*"[Title/Abstract] OR ("antibod*"[All Fields] AND "response*"[Title/Abstract]) OR "immune response*"[Title/Abstract] OR "cellular immunit*"[Title/Abstract] OR "t cell response*"[Title/Abstract] OR ("antibod*"[All Fields] AND "titer*"[Title/Abstract]) |

| | | | |
|---|---|---|---|
| | | "Spike Glycoprotein, Coronavirus"[Mesh] | "spike protein"[Title/Abstract] OR "anti sars cov 2 spike antibod*"[Title/Abstract] OR "s protein"[Title/Abstract] OR "spike protein antibod*"[Title/Abstract] OR "anti sars cov 2 spike protein antibod*"[Title/Abstract] OR "s protein antibod*"[Title/Abstract] OR "anti sars cov 2 igg"[Title/Abstract] OR "anti sars cov 2 quantivac elisa"[Title/Abstract] OR "anti spike antibod*"[Title/Abstract] OR "covid 19 antibod*"[Title/Abstract] OR "coronavirus spike protein"[Title/Abstract] OR "neutralising antibod*"[Title/Abstract] OR "neutralizing antibod*"[Title/Abstract] |
| | | "Safety"[Mesh] | "safety"[Title/Abstract] OR "side effect*"[Title/Abstract] OR "adverse effect*"[Title/Abstract] OR "adverse event*"[Title/Abstract] |
| | | "Vaccine Efficacy"[Mesh] | "efficacy"[Title/Abstract] OR "effectiveness"[Title/Abstract] |

***Table S2. Pubmed search strategy***

| Search no. | Query | Results |
|---|---|---|
| #1 | ("Adult"[Mesh] OR "Young Adult"[Mesh]) OR ("adult*"[Title/Abstract] OR "young adult"[Title/Abstract]) | 8,470,351 |
| #2 | ("Middle Aged"[Mesh]) OR ("middle age*"[Title/Abstract] OR "age"[Title/Abstract] OR "older adult*"[Title/Abstract]) | 6,306,623 |
| #3 | (#1) OR (#2) | 9,469,885 |
| #4 | ("COVID-19 Vaccines"[Mesh]) OR ("covid 19 vaccine*"[Title/Abstract] OR "coronavirus vaccine*"[Title/Abstract] OR "sars cov 2 vaccine*"[Title/Abstract] OR "ncov vaccine*"[Title/Abstract]) | 15,668 |
| #5 | ("Immunity"[MeSH Terms] OR "immunity, cellular"[MeSH Terms] OR "immunity, heterologous"[MeSH Terms] OR "immunity, humoral"[MeSH Terms]) OR ("immunit*"[Title/Abstract] OR "humoral immunit*"[Title/Abstract] OR ("antibod*"[All Fields] AND "response*"[Title/Abstract]) OR "immune response*"[Title/Abstract] OR "cellular immunit*"[Title/Abstract] OR "t cell response*"[Title/Abstract] OR ("antibod*"[All Fields] AND "titer*"[Title/Abstract])) | 895,807 |
| #6 | ("Spike Glycoprotein, Coronavirus"[Mesh]) OR ("spike protein"[Title/Abstract] OR "anti sars cov 2 spike antibod*"[Title/Abstract] OR "s protein"[Title/Abstract] OR "spike protein antibod*"[Title/Abstract] OR "anti sars cov 2 spike protein antibod*"[Title/Abstract] OR "s protein antibod*"[Title/Abstract] OR "anti sars cov 2 igg"[Title/Abstract] OR "anti sars cov 2 quantivac elisa"[Title/Abstract] OR "anti spike antibod*"[Title/Abstract] OR "covid 19 antibod*"[Title/Abstract] OR "coronavirus spike protein"[Title/Abstract] OR "neutralising antibod*"[Title/Abstract] OR "neutralizing antibod*"[Title/Abstract]) | 48,930 |
| #7 | ("Safety"[Mesh]) OR ("safety"[Title/Abstract] OR "side effect*"[Title/Abstract] OR "adverse effect*"[Title/Abstract] OR "adverse event*"[Title/Abstract]) | 1,144,576 |
| #8 | ("Vaccine Efficacy"[Mesh]) OR ("efficacy"[Title/Abstract] OR "effectiveness"[Title/Abstract]) | 1,401,732 |
| #9 | (((#5) OR (#6)) OR (#7)) OR (#8) | 3,015,497 |
| #10 | ((#3) AND (#4)) AND (#9) | 2,905 |

Limit: - none; last date of search- 19-04-22

*Table S3. Embase search strategy*

| Search no. | Queries | Results |
|---|---|---|
| #1 | 'adult'/exp OR 'adult' OR 'adult':ti,ab,kw OR 'adults':ti,ab,kw OR 'young adult'/exp OR 'young adult' OR 'middle aged'/exp OR 'middle aged' OR 'middle age':ti,ab,kw OR 'middle aged':ti,ab,kw | 10,780,252 |
| #2 | 'sars-cov-2 vaccine'/exp OR '2019-ncov vaccine' OR '2019-ncov virus vaccine' OR 'covid 19 vaccine' OR 'covid-19 mrna vaccine' OR 'covid-19 recombinant protein vaccine' OR 'covid-19 vaccine' OR 'covid-19 vaccines' OR 'covid-19 virus vaccine' OR 'covid19 vaccine' OR 'covid19 virus vaccine' OR 'hcov-19 vaccine' OR 'hcov-19 virus vaccine' OR 'human coronavirus 2019 vaccine' OR 'sars coronavirus 2 vaccine' OR 'sars-cov-2 inactivated vaccine' OR 'sars-cov-2 mrna vaccine' OR 'sars-cov-2 recombinant protein vaccine' OR 'sars-cov-2 vaccine' OR 'sars-cov-2 virus vaccine' OR 'sars2 vaccine' OR 'sars2 virus vaccine' OR 'wuhan coronavirus vaccine' OR 'coronavirus disease 2019 vaccine' OR 'inactivated covid-19 vaccine' OR 'inactivated sars-cov-2 vaccine' OR 'mrna covid-19 vaccine' OR 'mrna sars-cov-2 vaccine' OR 'ncov-2019 vaccine' OR 'ncov-2019 virus vaccine' OR 'novel 2019 coronavirus vaccine' OR 'novel coronavirus 2019 vaccine' OR 'severe acute respiratory syndrome 2 vaccine' OR 'severe acute respiratory syndrome coronavirus 2 vaccine' | 18,740 |
| #3 | 'vaccine immunogenicity'/exp OR 'immunogenicity, vaccine' OR 'vaccine immunogenecity' OR 'vaccine immunogenicity' OR 'coronavirus spike glycoprotein'/exp OR 'coronavirus s glycoprotein' OR 'coronavirus glycoprotein s' OR 'coronavirus spike glycoprotein' OR 'coronavirus spike glycoproteins' OR 'coronavirus spike protein' OR 'coronavirus spike proteins' OR 'spike glycoprotein, coronavirus' OR 'adverse event'/exp OR 'adverse effect' OR 'adverse effects' OR 'adverse event' OR 'adverse events' OR 'adverse reaction' OR 'side effect'/exp OR 'side effect' OR 'side reaction' OR 'vaccine'/exp OR 'combined vaccine' OR 'vaccin' OR 'vaccine' OR 'vaccine control' OR 'vaccine efficacy' OR 'vaccine potency' OR 'vaccine safety' OR 'vaccines' OR 'vaccines, combined' | 2,343,989 |
| #4 | #1 AND #2 AND #3 | 8,692 |

Limit: - none; last date of search- 18-04-22

*Table S4. Scopus search strategy*

| Search no. | Queries | Results |
|---|---|---|
| #1 | TITLE-ABS-KEY ( "adult*" OR "young adult" OR "middle age*" OR "age" ) | 11,647,399 |
| #2 | TITLE-ABS-KEY ( "COVID 19 vaccin*" OR "coronavirus vaccin*" OR "sars cov 2 vaccin*" OR "ncov vaccin*" ) | 19,774 |
| #3 | TITLE-ABS-KEY ( "immunit*" OR "humoral immunit*" OR "immune response*" OR "cellular immunit*" OR "t cell response*" ) OR ( ALL ( "antibod*" ) AND TITLE-ABS-KEY ( "response*" ) ) OR ( ALL ( "antibod*" ) AND TITLE-ABS-KEY ( "titer*" ) ) | 1,331,255 |
| #4 | TITLE-ABS-KEY ( ( "spike protein" OR "anti sars cov 2 spike antibod*" OR "s protein" OR "spike protein antibod*" OR "anti sars cov 2 spike protein antibod*" OR "s protein antibod*" OR "anti sars cov 2 igg" OR "anti sars cov 2 quantivac elisa" OR "anti spike antibod*" OR "covid 19 antibod*" OR "coronavirus spike protein" OR "neutralising antibod*" OR "neutralizing antibod*" ) ) | 69,519 |
| #5 | TITLE-ABS-KEY ( ( "safety" OR "side effect*" OR "adverse effect*" OR "adverse event*" ) ) | 2,719,991 |
| #6 | TITLE-ABS-KEY ( "efficacy" OR "effectiv*" ) | 7,854,548 |
| #7 | PUBYEAR > 2019 | 8,618,917 |
| #8 | #3 OR #4 OR #5 OR #6 | 10,713,607 |
| #9 | #1 AND #2 AND #7 AND #8 | 5,182 |

**Table S5. Web of Science search strategy**

| Search no. | Queries | Results |
|---|---|---|
| #1 | TS=(adult OR young adult OR middle age OR older adult) | 1,975,375 |
| #2 | TS=(COVID-19 vaccine OR sars-cov-2 vaccine) | 21,626 |
| #3 | TS=(immunogenicity OR immune* OR immune response OR antibody) | 1,648,927 |
| #4 | TS=(spike glycoprotein OR spike protein OR covid-19 antibod* OR neutralizing antibod* OR anti-sars-cov-2) | 79,281 |
| #5 | TS=(safety OR side effect OR adverse effect OR adverse event) | 1,843,421 |
| #6 | TS=(vaccine efficacy OR effectiveness) | 1,271,579 |
| #7 | #3 OR#4 OR #5 OR #6 | 4,517,221 |
| #8 | #1 AND #2 AND #7 | 1,195 |

TS = topic (search for title, abstracts, keywords)

Limit: - none; last date of search- 18-04-22

**Table S6. Cochrane search strategy**

Date Run: 19/04/2022 17:15:03

| Search | Queries | Results |
| --- | --- | --- |
| #1 | MeSH descriptor: [Adult] explode all trees | 490970 |
| #2 | MeSH descriptor: [Middle Aged] explode all trees | 332926 |
| #3 | #1 OR #2 | 490970 |
| #4 | MeSH descriptor: [COVID-19 Vaccines] explode all trees | 119 |
| #5 | MeSH descriptor: [Immunity, Humoral] explode all trees | 180 |
| #6 | MeSH descriptor: [Immunity, Cellular] explode all trees | 1727 |
| #7 | MeSH descriptor: [Immunity, Heterologous] explode all trees | 10 |
| #8 | MeSH descriptor: [Immunity] explode all trees | 4177 |
| #9 | MeSH descriptor: [Immunoglobulins] explode all trees | 28684 |
| #10 | MeSH descriptor: [Spike Glycoprotein, Coronavirus] explode all trees | 25 |
| #11 | MeSH descriptor: [Antibodies, Neutralizing] explode all trees | 539 |
| #12 | MeSH descriptor: [Safety] explode all trees | 4142 |
| #13 | MeSH descriptor: [Vaccine Efficacy] explode all trees | 12 |
| #14 | #5 OR #6 OR #7 OR #8 OR #9 OR #10 OR #11 OR #12 OR #13 | 34832 |
| #15 | #3 AND #4 AND #14 | 67 |

**Table S7. Search strategies in other databases**

| Serial no. | Databases | Search queries | Results |
|---|---|---|---|
| 1 | Science Direct | Searching strategy: "COVID-19 vaccine" AND ("effectiveness" OR "efficacy" OR "immunogenicity" OR "safety") AND "adult" | 134 |
| 2 | POPLINE | ((Abstract:(immune response* after COVID-19 vaccin*)) OR (Abstract:(spike protein antibod* post COVID-19 vaccin*)) OR (Abstract:(cellular immunity after SARS-CoV-2 vaccine*)) OR (SARS-CoV-2 antibod*) OR (neutralizing antibod*) OR (neutralising antibod*)) AND (Abstract:(COVID-19 vaccine* OR SARS-CoV-2 vaccine*)) AND (safety OR adverse effect* OR side effect*) AND (efficacy OR effective OR effectiveness) NOT (Abstract:(cancer)) NOT (Abstract:(leukemia)) NOT (Abstract:(hemodialysis)) NOT (Abstract:(animal*)) NOT (TitleCombined:(transplant*)) NOT (TitleCombined:(patient*)) NOT (TitleCombined:(veteran*)) NOT (TitleCombined:(adolescent children cancer leukemia hemodialysis thalassemia pregnant geriatric)) | 653 |
| 3 | HINARI | ((Abstract:(immune response* after COVID-19 vaccin*)) OR (Abstract:(spike protein antibod* post COVID-19 vaccin*)) OR (Abstract:(cellular immunity after SARS-CoV-2 vaccine*)) OR (SARS-CoV-2 antibod*) OR (neutralizing antibod*) OR (neutralising antibod*)) AND (Abstract:(COVID-19 vaccine* OR SARS-CoV-2 vaccine*)) AND (safety OR adverse effect* OR side effect*) AND (efficacy OR effective OR effectiveness) NOT (Abstract:(cancer)) NOT (Abstract:(leukemia)) NOT (Abstract:(hemodialysis)) NOT (Abstract:(animal*)) NOT (TitleCombined:(transplant*)) NOT (TitleCombined:(patient*)) NOT (TitleCombined:(veteran*)) NOT (TitleCombined:(adolescent children cancer leukemia hemodialysis thalassemia pregnant geriatric)) | 200 |
| 4 | Google Scholar | with all of the words- immune response post COVID 19 vaccination<br>with the exact phrase- COVID 19 vaccine<br>with at least one of the words- spike protein antibody OR COVID-19 vaccines<br>without the words- cancer leukemia hemodialysis malignancy adolescent rheumatoid arthritis | 520 |
| 5 | ClinicalTrials.gov | "COVID-19" and "vaccine". Filters: Status = "Completed" or "terminated" or "unknown status", Population = "Adult (18-64)". | 253 |
| 6 | WHO ICTRP | COVID-19 Special collection. Filtered using Excel:<br>1 - Last Refreshed on: 2020 or 2021 or 2022;<br>2 - Public title: contains "vaccin"<br>3 - Inclusion agemin: >= 18 yrs old, or NA/blank<br>4 - Inclusion agemax: <= 65 yrs old, or NA/blank | 584 |

(NEJM= COVID-19 vaccines = 97 results)
