## Supplementary File 2. GRADE evidence profile for "Comparative Immunogenicity, Safety and Efficacy Profiles of four COVID-19 Vaccine types in healthy adults: Systematic Review cum Meta-analysis of Clinical Trial data"

### Comparative Immunogenicity, Safety and Efficacy profiles of four COVID-19 Vaccine types in healthy adults: Systematic Review cum Meta-analysis of Clinical Trial data

Si Qi YOONG[a], Priyanka BHOWMIK[b], Debprasad DUTTA[c,d, *]

**Authors Affiliations:**

Si Qi YOONG

[a] Alice Lee Centre for Nursing Studies, Yong Yoo Lin School of Medicine

National University of Singapore, Singapore

Priyanka BHOWMIK

[b] Tbilisi State Medical University, Georgia

Debprasad DUTTA*

[c] Mazumdar Shaw Center for Translational Research (MSCTR), Mazumdar Shaw Medical Foundation, Bangalore, India

[d] Institute of Infection, Veterinary and Ecological Sciences (IVES), University of Liverpool, Liverpool, UK

| Certainty assessment | | | | | | | № of participants | | Effect | | Certainty of the evidence (GRADE)* |
|---|---|---|---|---|---|---|---|---|---|---|---|
| № of studies | Study design | Risk of bias | Inconsistency | Indirectness | Imprecision | Other considerations | Vaccine | Placebo | Relative (95% CI) | Absolute (95% CI) | |
| 7 days after first dose (local adverse events) | | | | | | | | | | | |
| 12 | randomised trials | not serious | not serious | not serious | not serious | none | 341/752 (45.3%) | 70/549 (12.8%) | RR 2.88 (1.78 to 4.67) | 240 more per 1,000 (from 99 more to 468 more) | ⊕⊕⊕⊕ High |
| 7 days after first dose (systemic adverse events) | | | | | | | | | | | |
| 10 | randomised trials | not serious | not serious | not serious | serious[a] | none | 243/673 (36.1%) | 123/471 (26.1%) | RR 1.30 (0.89 to 1.91) | 78 more per 1,000 (from 29 fewer to 238 more) | ⊕⊕⊕◯ Moderate |
| 7 days after second dose (local adverse events) | | | | | | | | | | | |
| 10 | randomised trials | not serious | not serious | not serious | not serious | none | 306/702 (43.6%) | 56/491 (11.4%) | RR 2.61 (1.38 to 4.90) | 184 more per 1,000 (from 43 more to 445 more) | ⊕⊕⊕⊕ High |
| 7 days after second dose (systemic adverse events) | | | | | | | | | | | |
| 7 | randomised trials | not serious | not serious | not serious | not serious | none | 234/600 (39.0%) | 80/405 (19.8%) | RR 2.24 (1.61 to 3.11) | 245 more per 1,000 (from 120 more to 417 more) | ⊕⊕⊕⊕ High |
| 1 month after first dose (any adverse events) | | | | | | | | | | | |
| 6 | randomised trials | not serious | serious[b] | not serious | serious[a] | none | 71/253 (28.1%) | 41/144 (28.5%) | RR 1.04 (0.66 to 1.65) | 11 more per 1,000 (from 97 fewer to 185 more) | ⊕⊕◯◯ Low |
| 1 month after second dose (any adverse events) | | | | | | | | | | | |
| 7 | randomised trials | not serious | not serious | not serious | serious[a] | none | 71/358 (19.8%) | 33/171 (19.3%) | RR 1.20 (0.83 to 1.73) | 39 more per 1,000 (from 33 fewer to 141 more) | ⊕⊕⊕◯ Moderate |

| | | | | | | | | | | | |
|---|---|---|---|---|---|---|---|---|---|---|---|
| Overall adverse events (7 days) | | | | | | | | | | | |
| 8 | randomised trials | not serious | serious[b] | not serious | not serious | none | 483/1028 (47.0%) | 122/575 (21.2%) | RR 1.68 (1.21 to 2.34) | 144 more per 1,000 (from 45 more to 284 more) | ⊕⊕⊕◯ Moderate |
| Overall adverse events (1 month) | | | | | | | | | | | |
| 9 | randomised trials | not serious | not serious | not serious | not serious | none | 380/1231 (30.9%) | 178/684 (26.0%) | RR 1.21 (1.01 to 1.45) | 55 more per 1,000 (from 3 more to 117 more) | ⊕⊕⊕⊕ High |
| 7 days after primary series (neutralising antibodies – live virus neutralisation assay) | | | | | | | | | | | |
| 4 | randomised trials | not serious | very serious[b] | not serious | not serious | strong association[c] | 191 | 90 | - | SMD 2.51 higher (1.58 higher to 3.44 higher) | ⊕⊕⊕◯ Moderate |
| 14 days after primary series (neutralising antibodies – live virus neutralisation assay) | | | | | | | | | | | |
| 14 | randomised trials | not serious | very serious[b] | not serious | not serious | publication bias strongly suspected strong association[c,d] | 882 | 527 | - | SMD 4.3 higher (3.54 higher to 5.07 higher) | ⊕⊕◯◯ Low |
| 28 days after primary series (neutralising antibodies – live virus neutralisation assay) | | | | | | | | | | | |
| 12 | randomised trials | not serious | very serious[b] | not serious | not serious | strong association[c] | 916 | 578 | - | SMD 4.7 higher (3.55 higher to 5.85 higher) | ⊕⊕⊕◯ Moderate |
| 28 days after primary series (neutralising antibodies – pseudovirus neutralisation assay) | | | | | | | | | | | |
| 6 | randomised trials | not serious | very serious[b] | not serious | not serious | strong association[c] | 339 | 248 | - | SMD 3.41 higher (2.48 higher to 4.34 higher) | ⊕⊕⊕◯ Moderate |
| 14 days after primary series (anti-RBD IgG) | | | | | | | | | | | |
| 10 | randomised trials | not serious | very serious[b] | not serious | not serious | strong association[c] | 694 | 436 | - | SMD 5.68 higher (3.95 higher to 7.42 higher) | ⊕⊕⊕◯ Moderate |
| 28 days after primary series (anti-RBD IgG) | | | | | | | | | | | |

| | | | | | | | | | | | |
|---|---|---|---|---|---|---|---|---|---|---|---|
| 13 | randomised trials | not serious | very serious[b] | not serious | not serious | strong association[c] | 1265 | 1081 | - | SMD 4.31 higher (3.21 higher to 5.42 higher) | ⊕⊕⊕◯ Moderate |
| 7 days after primary series (anti-S IgG) | | | | | | | | | | | |
| 3 | randomised trials | not serious | very serious[b] | not serious | not serious | strong association[c] | 110 | 88 | - | SMD 3.71 higher (1.01 higher to 6.42 higher) | ⊕⊕⊕◯ Moderate |
| 14 days after primary series (anti-S IgG) | | | | | | | | | | | |
| 9 | randomised trials | not serious | very serious[b] | not serious | not serious | strong association[c] | 1076 | 930 | - | SMD 5.48 higher (3.66 higher to 7.29 higher) | ⊕⊕⊕◯ Moderate |

**CI:** confidence interval; **RR:** risk ratio

*Explanations*

a. 95% CI crosses RR of 1.0

b. Heterogeneity was significant

c. SMD > 0.8 (large effect size)

d. Begg's and Egger's test was significant

***GRADE Working Group grades of evidence**

**High certainty:** we are very confident that the true effect lies close to that of the estimate of the effect.

**Moderate certainty:** we are moderately confident in the effect estimate: the true effect is likely to be close to the estimate of the effect, but there is a possibility that it is substantially different.

**Low certainty:** our confidence in the effect estimate is limited: the true effect may be substantially different from the estimate of the effect.

**Very low certainty:** we have very little confidence in the effect estimate: the true effect is likely to be substantially different from the estimate of effect.
