## Supplementary File 3. Supplementary meta-analysis figures for "Comparative Immunogenicity, Safety and Efficacy Profiles of four COVID-19 Vaccine types in healthy adults: Systematic Review cum Meta-analysis of Clinical Trial data"

### Comparative Immunogenicity, Safety and Efficacy profiles of four COVID-19 Vaccine types in healthy adults: Systematic Review cum Meta-analysis of Clinical Trial data

Si Qi YOONG[a], Priyanka BHOWMIK[b], Debprasad DUTTA[c,d, *]

**Authors Affiliations:**

**Authors Affiliations:**

Si Qi YOONG

[a] Alice Lee Centre for Nursing Studies, Yong Yoo Lin School of Medicine

National University of Singapore, Singapore

Priyanka BHOWMIK

[b] Tbilisi State Medical University, Georgia

Debprasad DUTTA*

[c] Mazumdar Shaw Center for Translational Research (MSCTR), Mazumdar Shaw Medical Foundation, Bangalore, India

[d] Institute of Infection, Veterinary and Ecological Sciences (IVES), University of Liverpool, Liverpool, UK

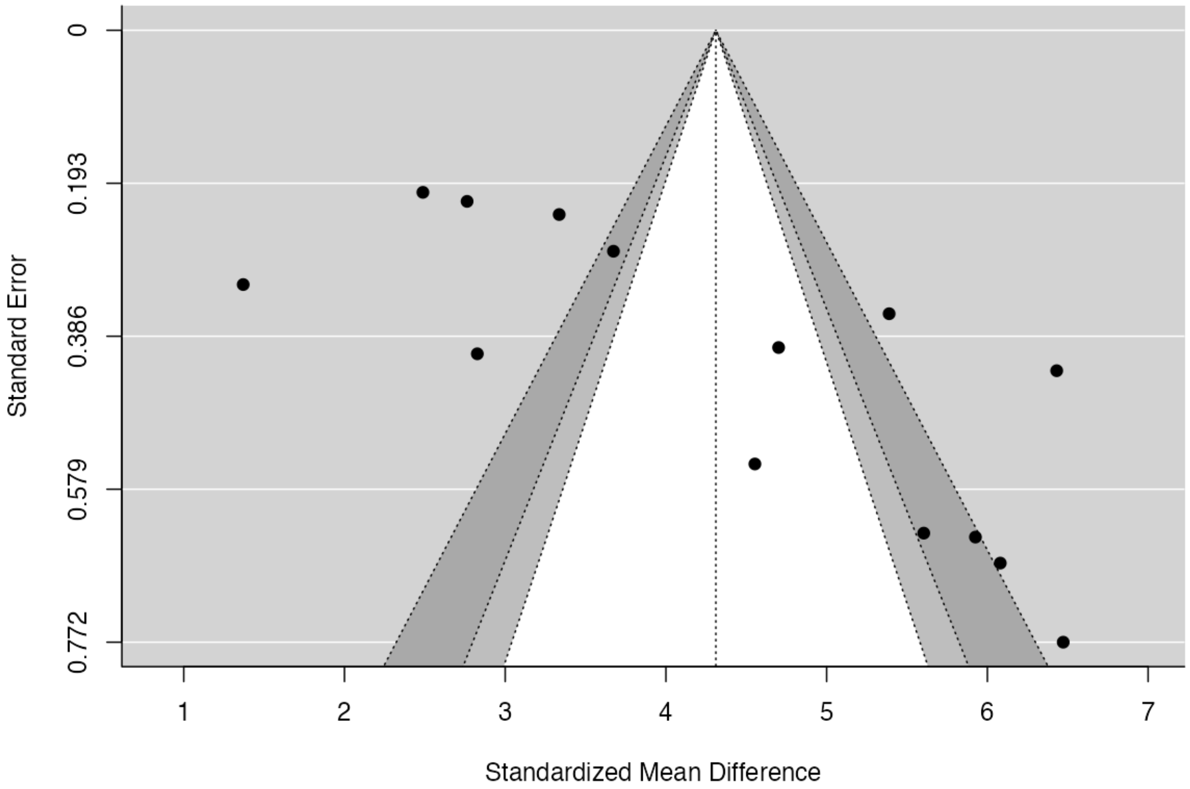


**Figure S1.** Funnel plot for publication bias assessment (with reference to Figure 3B. Log-transformed neutralizing antibody levels 14 days after COVID-19 vaccination, measured using live virus neutralization assays)

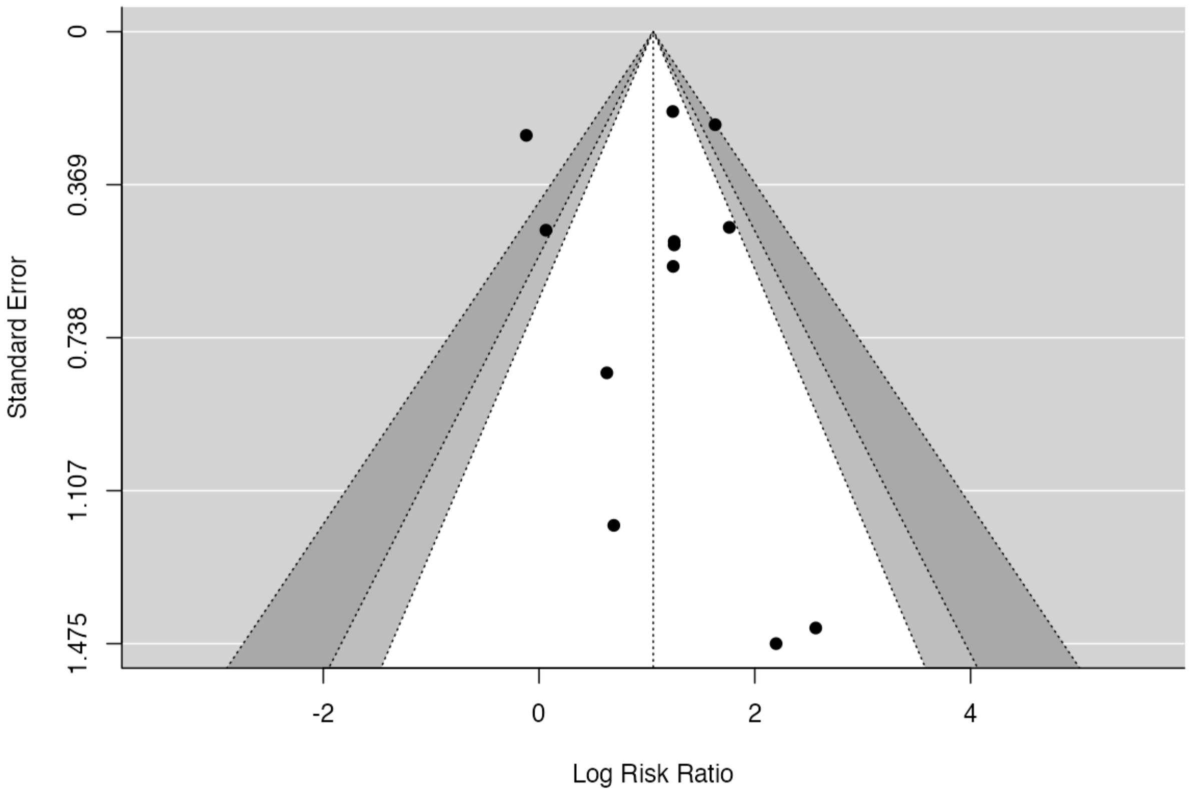


**Figure S2.** Funnel plot for publication bias assessment (with reference to Figure 5A. Local adverse events seven days after the first dose of a COVID-19 vaccine)

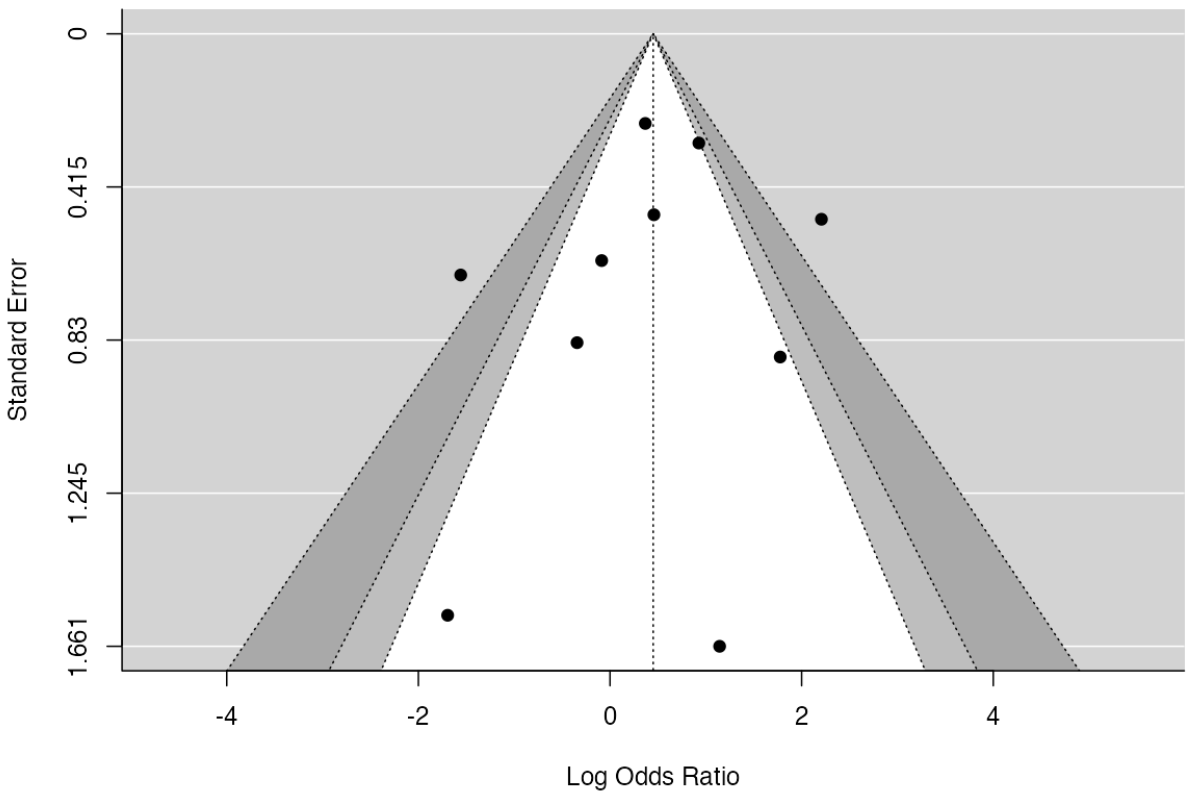


**Figure S3.** Funnel plot for publication bias assessment (with reference to Figure 5B. Systematic adverse events seven days after first dose of a COVID-19 vaccine)

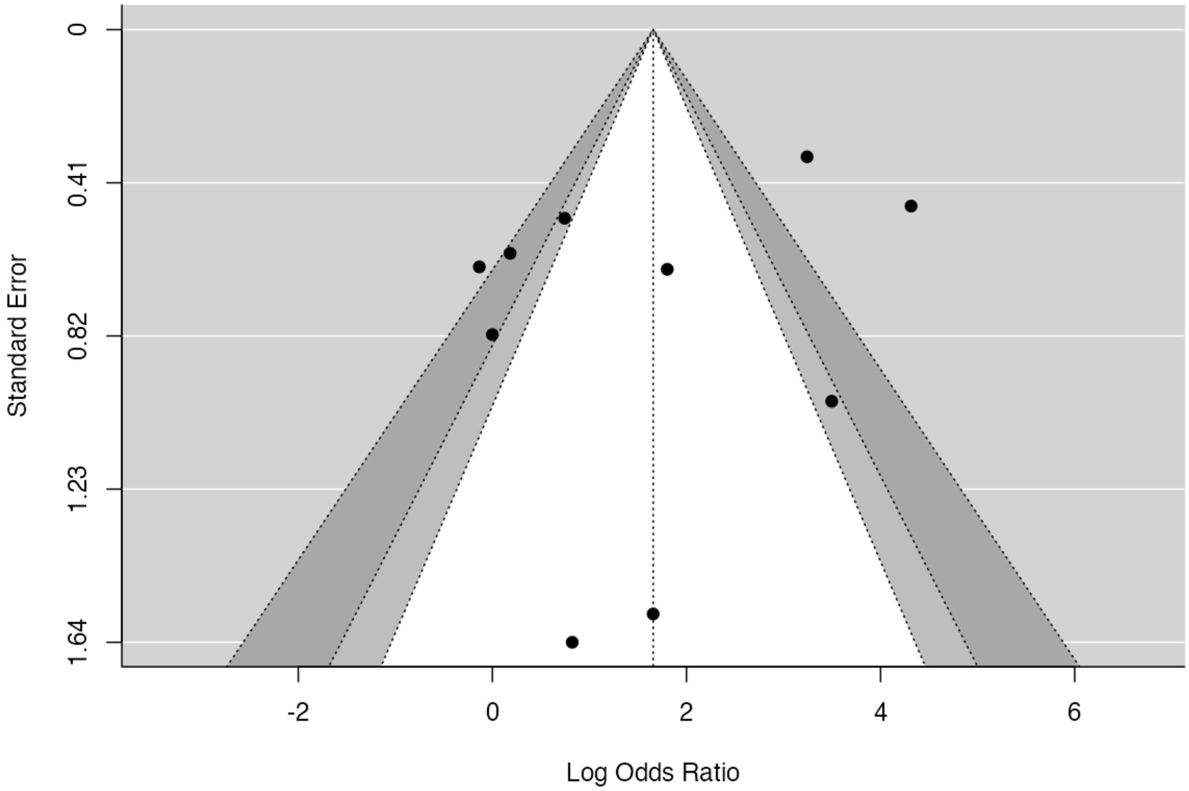


**Figure S4**. Funnel plot for publication bias assessment (with reference to Figure 6A. Local adverse events seven days after second dose of a COVID-19 vaccine)
